# Quantifying changes in vaccine coverage in mainstream media as a result of COVID-19 outbreak

**DOI:** 10.1101/2021.11.07.21266018

**Authors:** Bente Christensen, Daniel J Laydon, Tadeusz Chelkowski, Dariusz Jemielniak, Michaela Vollmer, Samir Bhatt, Konrad Krawczyk

**Affiliations:** Department of Mathematics and Computer Science, University of Southern Denmark, Odense, DK; Department of Infectious Disease Epidemiology, MRC Centre for Global Infectious Disease Analysis, Imperial College London, London, GB; Department of Management in the Network Society, Kozminski University, Warsaw, PL; Section of Epidemiology, Department of Public Health, University of Copenhagen, Copenhagen, DK

## Abstract

**Background:** Achieving vaccine-derived herd immunity depends on public acceptance of vaccination, which in turn relies on people’s understanding of its risks and benefits. The fundamental objective of public health messaging on vaccines is therefore the clear and concise communication of often complex information, and increasingly the countering of misinformation. The primary outlet shaping societal understanding is the mainstream online news media. There was widespread media coverage of the multiple vaccines that were rapidly developed in response to COVID-19. We studied vaccine coverage on the front pages of mainstream online news, using text-mining analysis to quantify the amount of information and sentiment polarization of vaccine coverage delivered to readers.

**Methods:** We analyzed 28 million articles from 172 major news sources, across 11 countries between July 2015 and April 2021. We employed keyword-based frequency analysis to estimate the proportion of coverage given to vaccines in our dataset. We performed topic detection using BERTopic and Named Entity Recognition to identify the leading subjects and actors mentioned in the context of vaccines. We used the Vader Python module to perform sentiment polarization quantification of all our English-language articles.

**Results:** We find that the proportion of headlines mentioning vaccines on the front pages of international major news sites increased from 0.1% to 3.8% with the outbreak of COVID-19. The absolute number of negatively polarized articles increased from a total of 6,698 before the COVID-19 outbreak 2015-2019 compared to 28,552 in 2020-2021. Overall, however, before the COVID-19 pandemic, vaccine coverage was slightly negatively polarized (57% negative) whereas with the outbreak, the coverage was primarily positively polarized (38% negative).

**Conclusions:** Because of COVID-19, vaccines have risen from a marginal topic to a widely discussed topic on the front pages of major news outlets. Despite a perceived rise in hesitancy, the mainstream online media, i.e. the primary information source to most individuals, has been strongly positive compared to pre-pandemic vaccine news, which was mainly negative. However, the pandemic was accompanied with an order of magnitude increase in vaccine news volume that due to pre-pandemic low frequency sampling bias may contribute to a perceived negative sentiment. These results highlight the important interactions between the volume of news and overall polarisation. To the best of our knowledge, our work is the first systematic text mining study of vaccines in the context of COVID-19.

## 1. Introduction

Theoretical models suggest that the herd immunity threshold for SARS-COV-2 requires two thirds of the population to be immunized, through either natural infection or vaccination. (1). Though multiple safe and effective vaccines were developed against SARS-COV-2 (2–4), one significant challenge in achieving pandemic control would be insufficient vaccine acceptance, or ‘vaccine hesitancy’ (5).

Vaccine hesitancy extends beyond COVID-19 as according to the World Health Organization (WHO), it is one of the ten biggest threats to global health. At its core, vaccine hesitancy is an issue of perception, rooted in the information individuals receive (6).

One of the main sources of vaccine information and misinformation is social media. Recent analysis of vaccine-related Tweets indicated that they are predominantly positively polarized (7). However, there is also substantial misinformation, including possibly coordinated efforts (8), which may affect attitudes (9). Another study showed that the volume of tweeted fake news in a country negatively correlates with its vaccine uptake (10). Anti-vaccination supporters on Twitter, share more conspiracy theories and make greater use of emotional language than pro-vaccination supporters (11). Moreover, vaccine discourse is highly politicized (12), and the likelihood of endorsing misinformation is ideologically driven (13,14).

Vaccine discourse is also about different objective values. Defending vaccines prioritizes community, while vaccine hesitancy pages focus on freedom (15). Another study showed a high proportion of parents’ opinions of vaccines expressed online were aggressive, accusatory or inaccurate, suggesting that blog comments are less reliable (16).

Despite substantial anti-vaccination content on social media, major news outlets play an important platform both for endorsement and discouragement of vaccines (17)(18). To the best of our knowledge, there are no systematic text mining studies of vaccines in the context of COVID-19 to date.

Here, we analyze online news media coverage of COVID-19 vaccines. We use text mining analysis to estimate the volume of online vaccine news coverage: i) before the COVID-19 pandemic; ii) before vaccine rollout, and iii) during vaccine rollout. We used ∼28 million front page headlines, collected from 11 different countries with robust online media. Because sentiment towards vaccines is influenced by the context in which they are mentioned, the most frequently mentioned topics in the different periods are gathered along with the most frequently mentioned companies and organizations. Our text mining study will hopefully inform future public health and vaccine communication, and possibly mitigate against vaccine hesitancy.

## 2. Methods

### 2.1 Curation of frontpage news articles database

We analysed the landing pages from major Online News Sources (ONS) in countries with a robust media presence. The data is fully described in a previous study (19), which focused on front-page news from 172 leading ONSs in 11 countries: Australia, Canada, France, Germany, Ireland, Italy, New Zealand, Russia, Spain, UK, USA and an international category. The data used articles from July 2015 to April 2021, that is, before COVID-19, during the COVID-19 pandemic but before vaccine rollout and during vaccine rollout. The updated dataset collected a total of 28,709,060 headlines, from which 14,638,278 were English-language and 14,070,782 were non-English-language.

### 2.2 Identifying vaccine headlines

Keywords for vaccines were used to identify whether a given headline was vaccine related or not. For non-English headlines, keywords were supplied by experts, who speak the individual languages. For the English headlines, we supplied keywords ourselves. The used keywords can be found in Table 1.

**Table 1:**
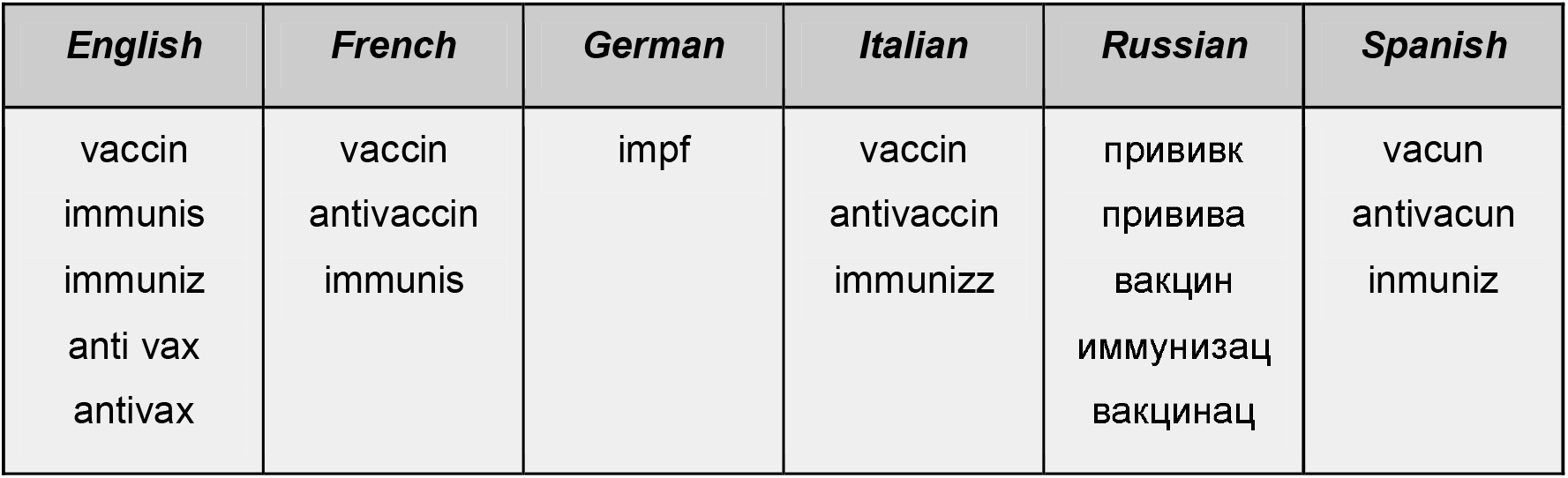
Keywords used to identify the vaccine headlines. The words are stemmed to capture the equivalence class of different forms of words (e.g. in German Impfung, impfen, Impfgegner etc. all map to impf).

All non-English headlines were stemmed using SnowballStemmer and case-folded, and they are presented as the word stems in Table 1. The English headlines were lemmatized using TreeTagger and all words were case-folded and punctuation removed, whereby words connected by a hyphen were separated into two words. English headlines were lemmatized to avoid misclassifications, e.g. “immunity” understood in a legal rather than biomedical sense.

To identify vaccine headlines, different techniques were used for different languages. In French, Italian, Russian and Spanish, titles and descriptions were tokenized, and if either contained at least one keyword, the headline was labelled as a vaccine headline. In English and German, the titles and descriptions were kept as strings, and a search was performed for keyword patterns. If one was present, the headline was assigned as a vaccine headline. This approach was chosen as all words containing one of the keyword patterns are vaccine related words. This search for patterns in the English headlines, along with the stemming challenge with immunisation is the reason that the English keywords given in Table 1 are shortenings, which in some cases are not real words nor stems.

### 2.3 Splitting the data into three vaccination-specific periods

We divided the data into three periods: i) the pre-COVID-19 era, ii) the COVID-19 pandemic pre-vaccine rollout and iii) the COVID-19 pandemic during vaccine rollout. The division of the data was based on clear changes within the media coverage with respect to vaccines and the coronavirus. On 9 January 2020 daily media coverage of the coronavirus began, and so this was chosen as the end of the pre-COVID-19 era. In the beginning of November 2020 there was a large increase in the relative media coverage of vaccines. This increase was based on the report published by Pfizer and BioNTech stating that their vaccine is 90 percent efficacious. November 9th 2020 was chosen as the cut-off date between pre-vaccine rollout and during-vaccine rollout. This resulted in the following three periods:

- Before COVID-19: July 2015 - 8 January, 2020
- COVID-19 before vaccine: 9 January, 2020 - 9 November, 2020
- COVID-19 with vaccine: 10 November, 2020 - 2 April, 2021

To identify changes in each period, the relative frequency of vaccines mentioned in the full dataset, along with the relative frequency of headlines containing either “COVID-19” or “coronavirus” was calculated at weekly intervals, using equation 1.

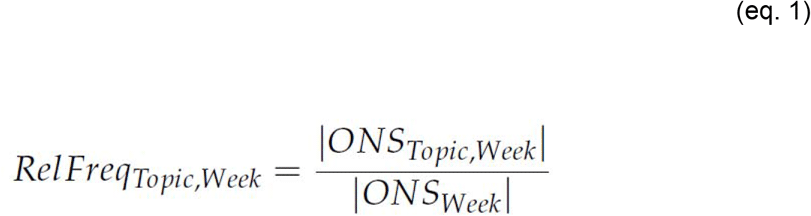

where |*ONS*_*Topic,Week*_ | is the number of headlines on a particular topic in a given week, and |*ONS*_*Week*_ | is the number of headlines in that same given week. The relative frequency was calculated with respect to two different topics, the first topic was vaccine where all headlines relating to vaccines were counted within the given topic. In the second case the relative frequency was calculated with respect to COVID-19, in this case all headlines containing either the keyword “coronavirus” or “COVID-19” was deemed within the given topic.

### 2.4 Topic Detection of the vaccine headlines in the three periods using BERTopic

Topics were detected for 91 English ONSs using BERTopic. Topics were not identified for the non-English ONSs, as finding the optimal number of topics within non-English ONSs would require languages to be handled separately, and would also require in-depth knowledge about each language. The BERTopic model was employed as it was previously successful in heterogeneous text mining (20,21) and it offers multiple pre-trained models. Additionally, scatter plots of the embeddings of the data from the three periods did not show a clear clustering of the headlines, which rules out different Topic Detection techniques (*please see supplementary figures 1, 2 and 3)*

To remove unwanted patterns from the text input to BERTopic, which could otherwise affect the model, all abbreviations, links and names referring to the different newspapers were removed. Additionally the word “news” was removed, along with words containing “immuniz”, “immunis” and “vaccin”, which were used to extract the vaccine headlines. The phrases “anti vax” and “antivax” were retained, as they refer to resistance towards vaccination.

Text input to BERTopic was normalized to reduce word variation. The headlines were lemmatized using TreeTagger combined with case-folding, where TreeTagger is a tool for annotating text with part-of-speech and lemma information, using a Markow tagger, which uses a decision tree to get reliable estimates. TreeTagger was also used to remove filler-words from headlines by only using words tagged as either a noun (including proper nouns), a verb or an adjective, removing words which contained little information about topics.

We employed a two-step evaluation method to identify the number of clusters reflecting the most common topics (Supplementary Section 1). The pseudo-code for this is illustrated in Algorithm 1. This method will in the remainder of this article be referred to as the *“Two Step Evaluation Method”* or *“TSEM”*.

#### Algorithm 1: The Two Step Evaluation Method (TSEM)

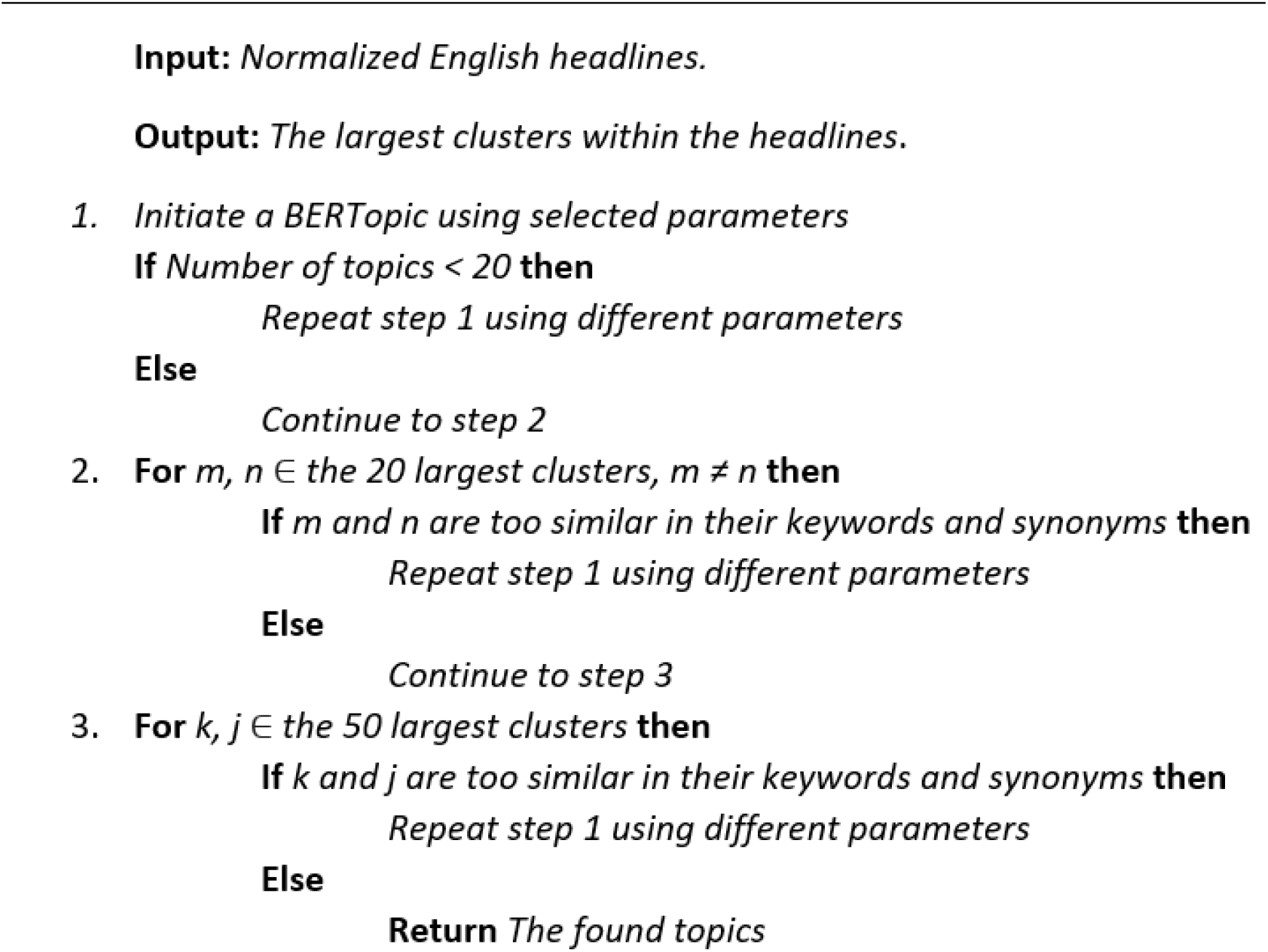

### 2.5 Named Entity Recognition of vaccine headlines using SpaCy

Named Entity Recognition (NER) identifies and categorizes words or strings of words for an entity, where an entity can be the name of a person, organization, location, work of art etc. We used NER to determine the companies and organizations that were mentioned frequently in the context of vaccination. This was performed on both the English and the non-English data using SpaCy, with different pipelines depending on the language. The pipelines were chosen according to the reported accuracy by SpaCy. In all cases the most accurate pipeline was used, which were en_core_web_trf, de_core_news_lg, fr_core_news_lg, it_core_news_lg, ru_core_news_lg and es_core_news_lg, where the two first letters in each pipeline refers to the language it was trained for.

Entities such as “*AstraZeneca-Oxford”* or “*Pfizer-BioNTech”* were split to count as separate entities. The occurrences of *“Johnson and Johnson”* and *“J&J”* were altered to *“Johnson & Johnson”*.

Counting individual entities was done on case-folded entities. Two bar plots were made, one containing the 30 most frequently occurring named entities from English ONSs, as well as one containing the 30 most frequently named entities from non-English ONSs.

### 2.6 Frequent n-grams with respect to the different vaccine manufacturers

To learn if a change in sentiment towards vaccination occured between the periods *COVID-19 before vaccine* to *COVID-19 with vaccine*, and whether such a change was caused by certain developments or changes over time, an assessment of the seven frequently occurring vaccine manufacturers found using NER was carried out. A dataset containing English headlines for each vaccine manufacturer was created. These were then assessed with respect to frequent bigrams and trigrams (referred to as n-grams henceforth). The lemmatized headlines created for the topic detection were used for this purpose.

For all vaccines and periods, the 50 most frequent n-grams were assessed. In some cases a combination of two bigrams, with almost the same count as a trigram, would combine to give that trigram. For instance the bigrams *(food, drug)* and *(drug, administr)* combined give the trigram *(food drug administr)*, where the small difference in count between bigrams and trigrams were caused, by *“Food and Drug Administration”* in some cases were referred to as *“Food and Drug Authority”* or *“Food and Drug Association”*. Based on this, such bigrams were removed, only keeping the trigrams. This was the case for *“Food and Drug Administration”, “Centers for Disease Control”* and *“European Medicines Agency”*. Additionally *“FDA”, “CDC”, “NIH”, “WHO”* and *“EMA”* were frequently occurring abbreviations among the frequent words with respect to some of the vaccines, whereby these were added to the number of occurrences of *“Food and Drug Administration”, “Center for Disease Control”, “National Institute of Health”, “World Health Organization”* and *“European Medicines Authority”* respectively. Other abbreviations such as *“NHS”, “HHS”, “PHE”* etc. were assessed with respect to frequent bigrams and trigrams, to learn if they could alter the occurrence of the most frequent ones, which was not the case. Likewise all bigrams were assessed and if they occurred the same number of times as a trigram containing the bigram, the bigram was removed. This was not the case if a bigram occurred more frequently than the trigram, whereby they both were kept. The bigram (european, union) was very frequent in most of the datasets, whereby this was removed.

### 2.7 Sentiment analysis of the vaccine headlines of three periods using Vader

Sentiment analysis was performed on all English headlines using VADER (22). Before assessing sentiment values, each headline’s raw score was calculated using the positive and negative sentiment values found using equation 2.

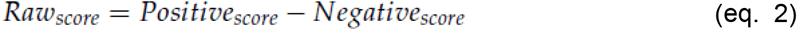

Some ONSs contained headlines with more negative or positive sentiment polarity than other ONSs, and furthermore this overall sentiment polarization could change over time. Therefore a comparison of sentiment towards vaccines between the periods and ONSs on the raw sentiment values would not show if a change in sentiment towards vaccines was due to an overall change in sentiment, or instead due to a change in sentiment with respect to vaccines. Therefore, to enable comparison of the periods and between the ONSs, each sentiment value for a vaccine headline was adjusted according to the overall average sentiment value. The adjustment was done using the VADER sentiment values (either raw or compound, denoted *S*_*ONS,Topic,Period*_) subtracting the mean sentiment value for the same ONS, with respect to non-vaccine headlines in the same period (either raw or compound) (denoted by *µ*_*ONS*,<u>*Topic*</u>,*Period*_). This is referred to as the Relative Sentiment Skew (RSS), and is given in equation 3

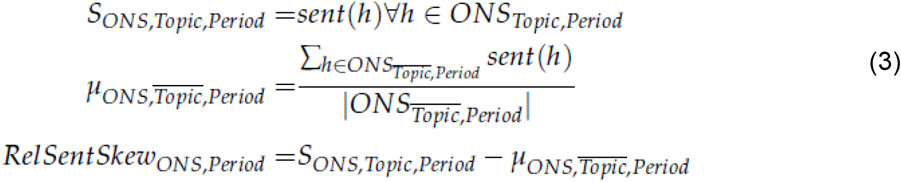

where *ONS*_*Topic,Period*_ is the collection of headlines of a given topic, for a given ONS in a specific period, *ONS*_<u>*Topic*</u>,*Period*_ is the collection of headlines not in the topic for that same ONS in all periods, *h* is a single headline, *sent*(*h*) is the sentiment value of h, while |*ONS*_<u>*Topic*</u>,*Period*_ | is the number of headlines not in the given topic for that same ONS in all periods. In this case the topic given in the above equation is vaccines. The raw scores were used to RSS for each headline, with respect to the three periods. These were illustrated in line plots, where the cumulative frequency showed the proportion of negative and positive RSS values of a certain value of smaller.

## 3. Results

### 3.1 Large increase in ratio of vaccine headlines with the rollout of COVID-19 vaccines

We calculated the percentage of vaccine coverage for each week within the entire period of data collection, plotted in Figure 1. This illustrates that before the COVID-19 outbreak, the ratio of vaccine headlines was relatively low (0.1% across our 172 ONSs). With the COVID-19 outbreak in early 2020, the ratio of vaccine headlines increased to be between one and two percent of the total amount of headlines each week.

**Figure 1:**
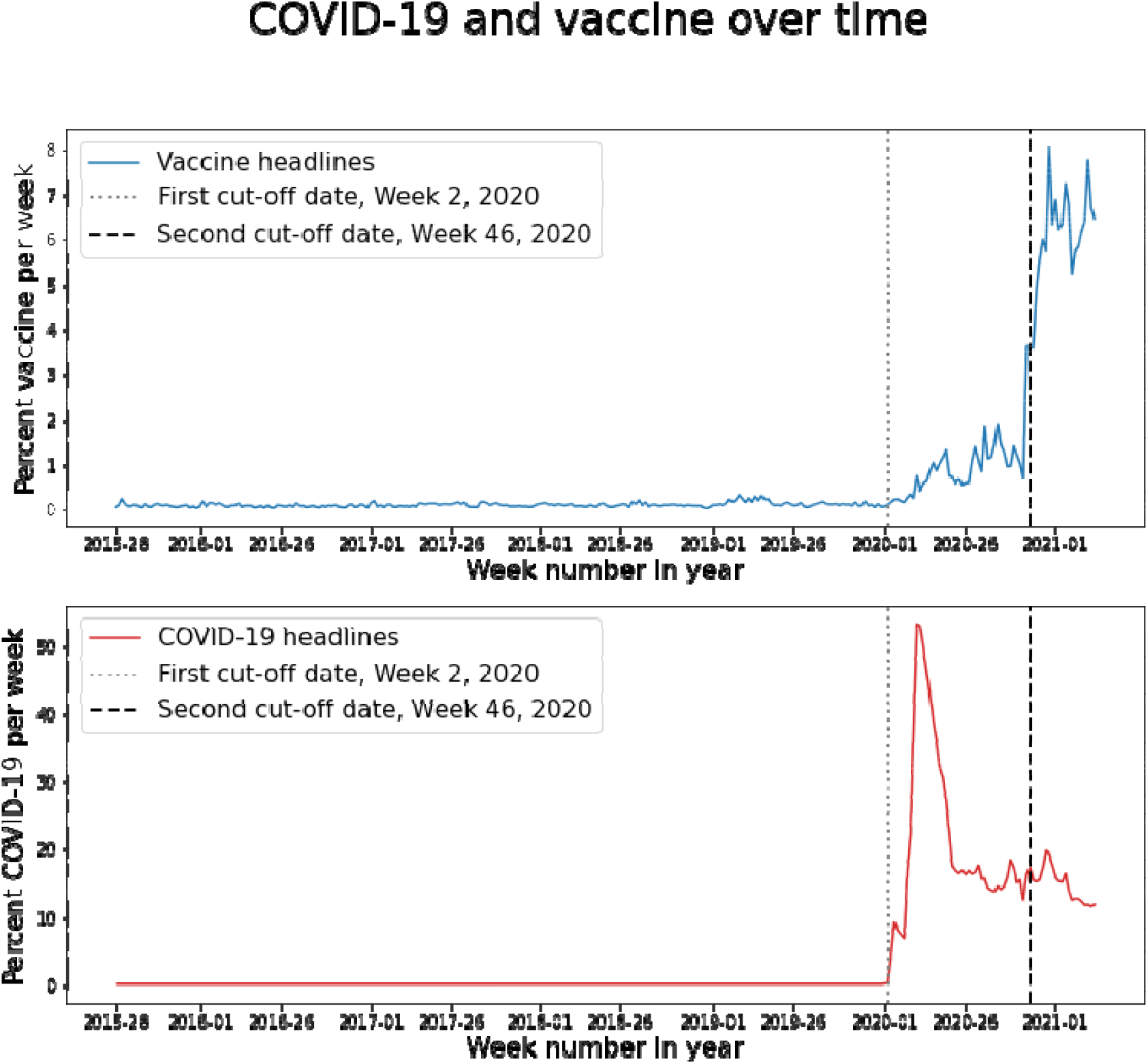
Percentage of headlines mentioning vaccines and COVID-19 in mainstream media. **Top**: The development in percentage of headlines containing vaccines over time, with an indicator of the first and second cut-off date. **Bottom:** The percentage of headlines containing “COVID-19” or “coronavirus” over time, with an indicator of the first and second cut-off date.

Our topic modelling indicates that the increased reporting on vaccines in this period was due to COVID-19 reporting. The ten most common topicsin vaccine coverage in the three periods as modelled by our BERTropic are shown in Table 2. The most common vaccine related topics following the outbreak of COVID-19 (i.e. during the second and third periods) are related to the pandemic rather than pre-COVID-19 vaccine related topics. Though this indicates that COVID-19 increased the frequency of vaccine coverage, the frequency of COVID-19 coverage is not directly correlated with that of vaccine coverage (Figure 1).

**Table 2:**
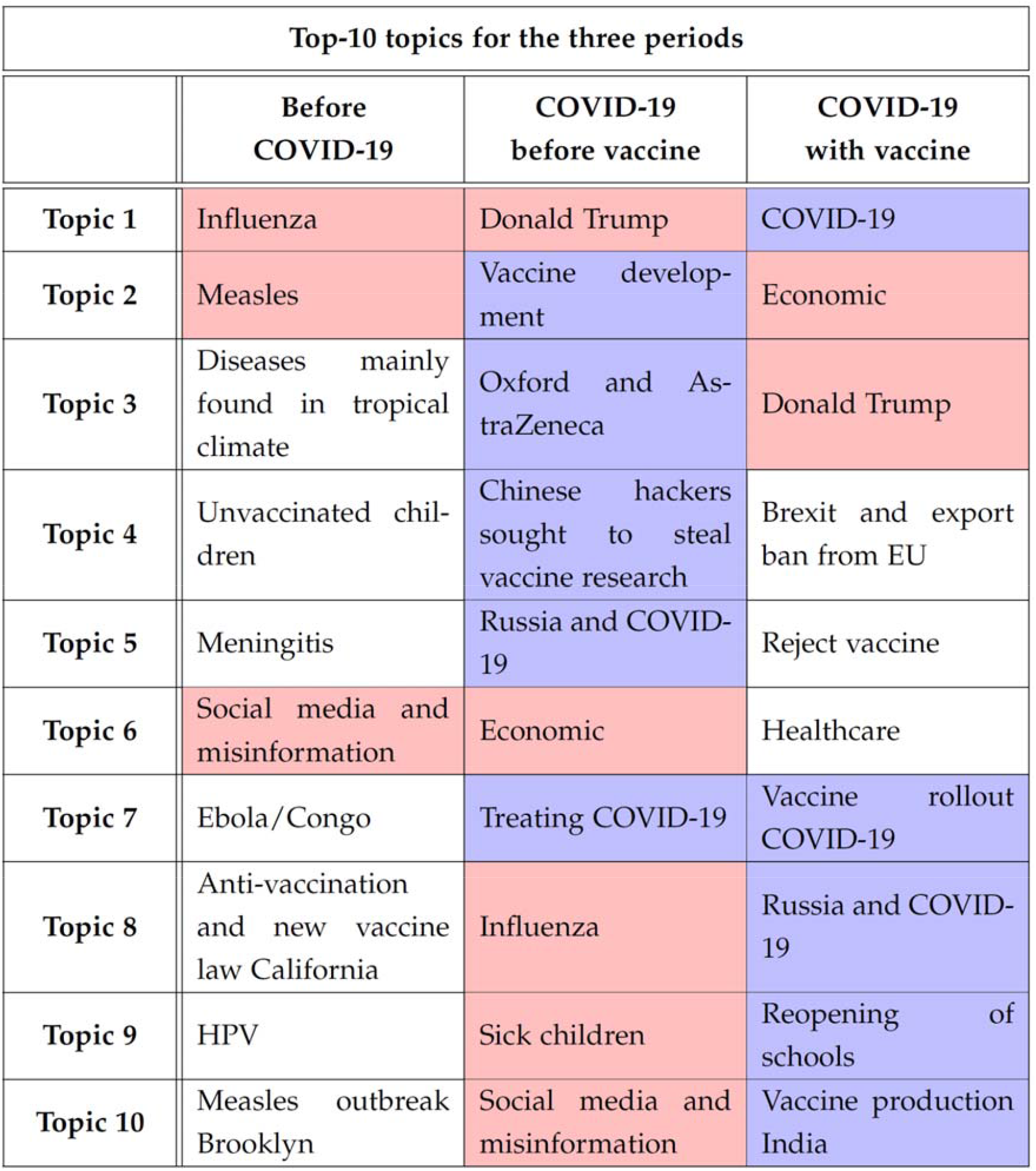
The ten most common topics from vaccine-related articles in the three periods in our study. The blue cells illustrate topics, which are directly COVID-19 related, while the red ones illustrate topics, which occur in more than one period.

Rather than dropping to a stable level like COVID-19 (Figure 1), the proportion of vaccine headlines increased from week 45 to 47 of 2020 to between six and eight percent and remained at this level until our cutoff date of 2nd April 2021. This increase is linked to the Pfizer and BioNTech press release on 9 November 2020, which reported that their vaccine has a 90 percent efficiency, paving the way for the rollout in the UK on the 2nd December 2020.

The relative frequencies of vaccine headlines were calculated for each period and each country, along with the international category (Figure 2). Figure 2 shows that the relative frequencies for different countries between each of the three periods show that the individual countries follow similar patterns with very limited attention towards vaccines in the period *“Before COVID-19”* and a steep rise after the introduction of the first SARS-CoV2 vaccine. These results illustrate that COVID-19 brought vaccine reporting from relatively rare coverage to the center of attention.

**Figure 2:**
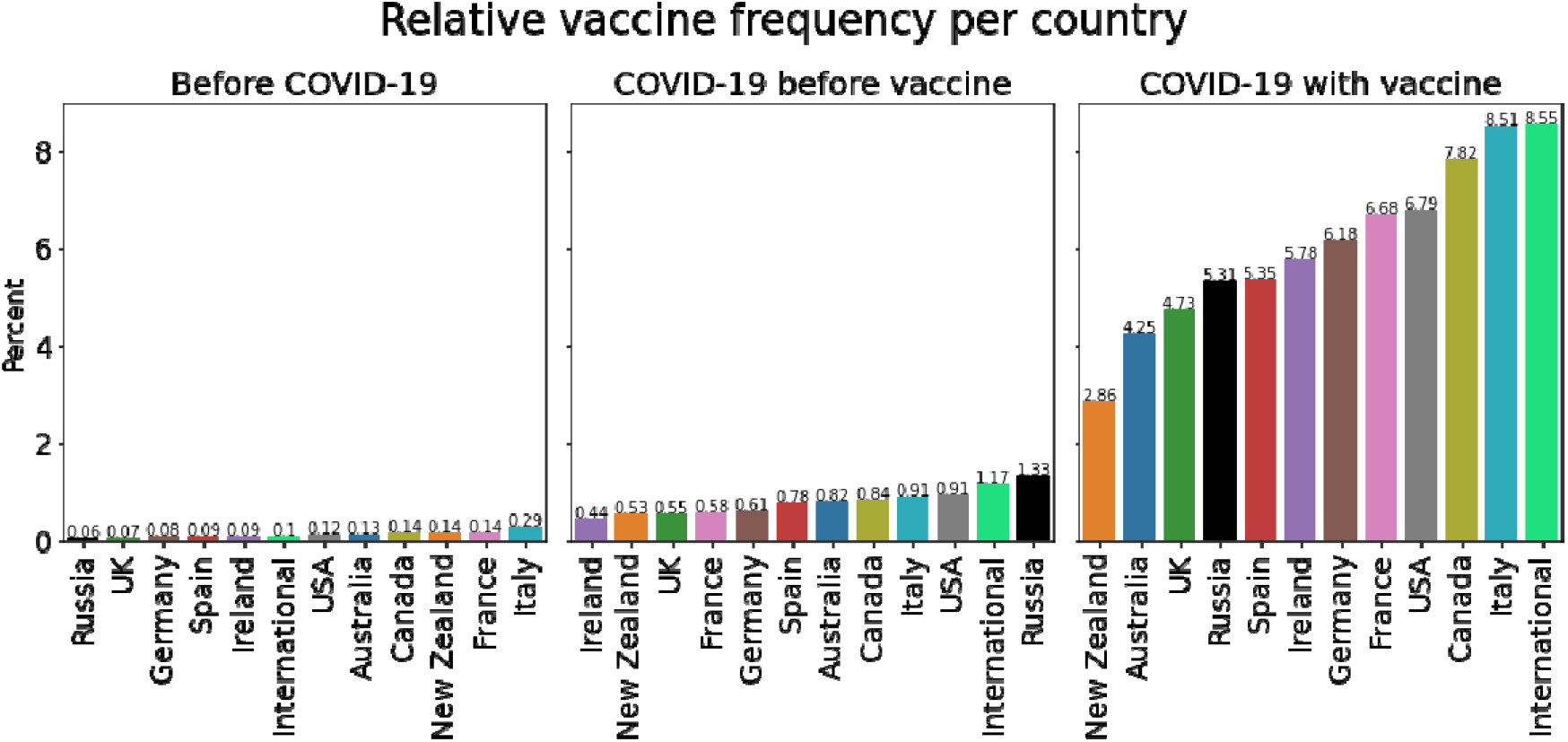
Relative vaccine frequency for each country including the international ONSs for each of the three periods.

### 3.2 Majority positive sentiment polarization of vaccine reporting with the outbreak of COVID-19 as opposed to the pre-pandemic era

Figure 3 shows VADER sentiment scores for text associated with vaccines in each period. The increased frequency of vaccine reporting during the pandemic led to an increase in the absolute number of negatively polarized articles from 6,698 in the period 2015-2019 to 28,552 in the shorter period 2020-2021. Overall however, polarization during the pandemic was majority positive (38.5% negatively polarized) as opposed to the pre-pandemic period where 57.1% of articles were negatively polarized. Table 2 demonstrates that the difference in sentiment between pre-COVID-19 and post-COVID-19 vaccine coverage can be explained by COVID-19 coverage. To investigate the difference in sentiment distribution between the two periods during the pandemic, we contrasted the topics and Named Entities mentioned in both periods.

**Figure 3.**
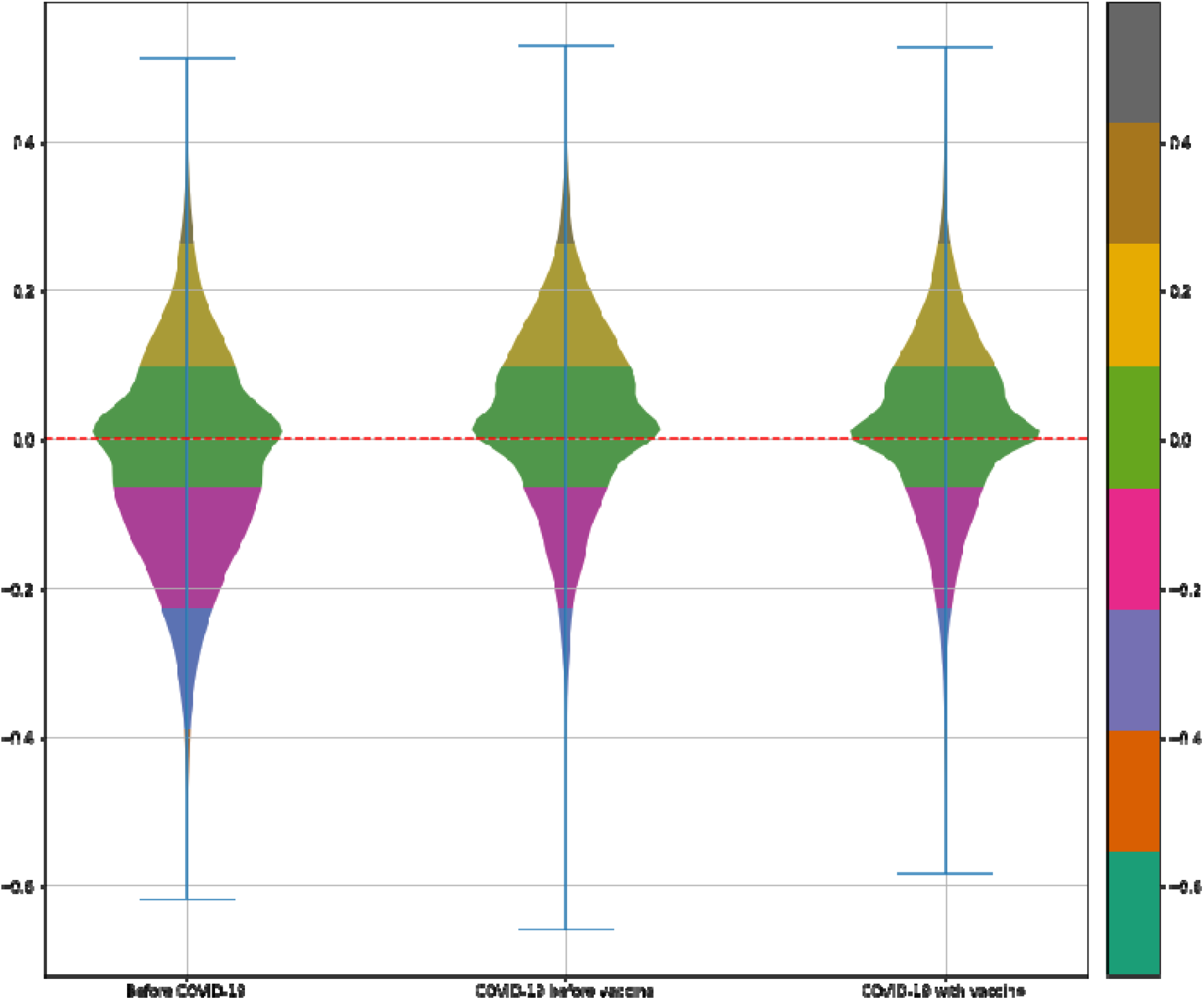
Relative Sentiment Skew (y-axis) of vaccine coverage in the three periods used in this study.

The period *“COVID-19 before vaccine”* can largely be interpreted as the period where the vaccines were developed, while *“COVID-19 with vaccine”* is the period where some vaccines were being rolled out, while others continue to be developed. Two topics *“Vaccine development”* and *“Vaccine rollout”* were extracted from the data giving two dataset with approximately the same size, namely: *“Vaccine development”* contained 846 headlines and *“Vaccine rollout”* contained 814 headlines.

Sentiment polarization was assessed with respect to the above two topics. RSS for the raw VADER sentiment with respect to the two topics is illustrated in Figure 4, which shows a change in sentiment towards the vaccines from the development and trial phase to the actual rollout of the vaccines. Figure 4 illustrates that for vaccine development almost the entire interquartile range is situated above the zero line. 22.8% of the headlines in *“Vaccine development”* had negative RSS, while 77.2% had positive RSS. This is very different in *“Vaccine rollout”*, where 66.09 percent had negative RSS, while 33.91 percent had positive RSS. Additionally, the widest area for *“Vaccine development”* lies above zero, while it lies below zero for *“Vaccine rollout”*. This means that RSS with highest frequency is positive with respect to *“Vaccine development”*, while it is negative for *“Vaccine rollout”*. The largest and smallest RSS for the two topics are quite different, with *“Vaccine Development”* lying in the range from approximately −0.3 to just below 0.5, while *“Vaccine rollout”* lies in the range from approximately −0.5 to 0.3, whereby their RSS values are equally spread, but their range are differently situated. This suggests that the difference in distributions of sentiments between the two COVID-19 periods could be attributed to more negative coverage during the vaccine rollout.

**Figure 4:**
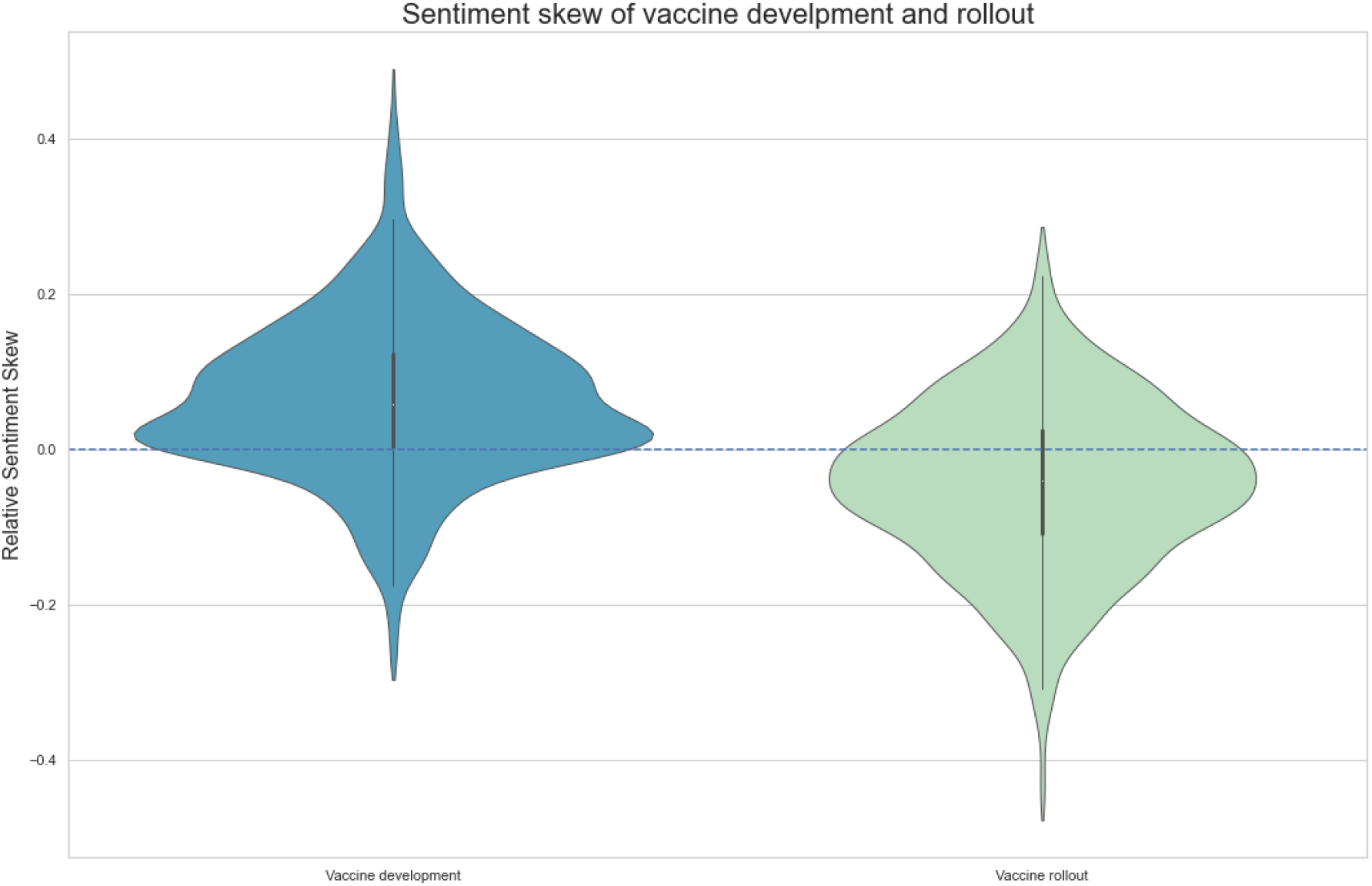
Relative Sentiment Skew for the topics “Vaccine development” and “Vaccine rollout” using the raw sentiment value.

### 3.3 The most common organizations mentioned in the context of COVID-19 vaccines and sentiment towards them

To gain more granular insight into the sentiment polarization of vaccine coverage in the pandemic period, we investigated the top entities mentioned in their context. For this task we employed Spacy to perform Named Entity Recognition. The 30 most frequent companies or organizations mentioned in vaccine context for the entire dataset, namely all three periods are illustrated in Figure 5.

**Figure 5:**
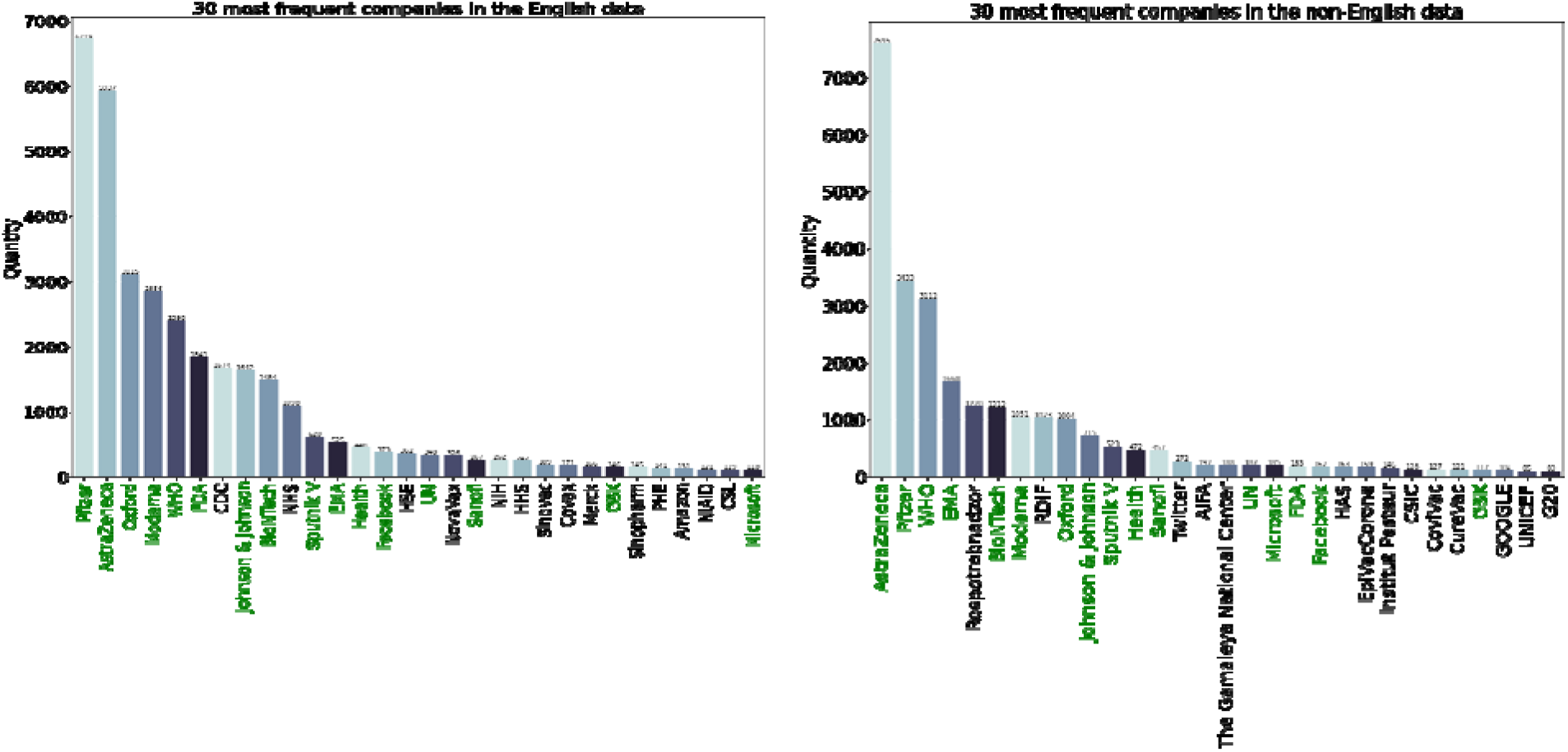
The 30 most frequent entities found in the English and non-English data, with respect to companies and organizations, where the green names are the organizations and companies which the English and non-English data have in common.

Unsurprisingly, the most common associations are with well known COVID-19 vaccine manufacturers, namely: *“AstraZeneca” (in collaboration with Oxford), “Pfizer” (in collaboration with BionTech), “BioNtech”, “Moderna”, “Oxford”, “Johnson & Johnson”* and *“Sputnik V”*, which all are found between the top 11 most frequent entities in both the English and non-English headlines. Though AtraZeneca & Oxford as well as Pfizer & BioNtech developed the vaccines as a partnership, they were frequently mentioned separately and thus we decided to keep them as separate entities.

Of the 30 most frequently occurring named entities, in the English and non-English headlines, there are 16 that occur in both datasets, colored green in Figure 5. The difference can be mainly attributed to National organizations or companies. For instance, *“NHS”* and *“HSE”*, are the health services in the UK and USA respectively and are solely found amongst the 30 most frequent English entities. *“Rospotrebnadzor”* is the Federal Service for Surveillance on Consumer Rights in Russia, and *“RDIF”* and *“PAH”* can also be linked to Russia and are found solely amongst the 30 most frequent non-English entities. Additionally, company names are the same across different languages, while this is not the case for some national organizations, for instance the abbreviation of World Health Organisation is in English WHO, while in French it is OMS.

The number of times that vaccine manufacturers are mentioned within headlines increased from negligible before COVID-19, to most frequently mentioned within the period of vaccine rollout (Table 3). With almost zero occurence of vaccine manufacturers before COVID-19, there is no foundation for further analysis within this period, and so the vaccine manufacturers are assessed only within the two periods of the COVID-19 outbreak.

**Table 3:** The 21 different subsets created with respect to the different vaccines and periods, including the number of headlines in each subset (seven vaccine manufacturers in three periods).

| The 21 created subset and their number of headlines |  |  |  |
| --- | --- | --- | --- |
|  | Before COVID-19 | COVID-19 before vaccine | COVID-19 with vaccine |
| AstraZeneca | 3 | 747 | 5,134 |
| BioNTech | 1 | 163 | 2,118 |
| Johnson & Johnson | 17 | 332 | 1,050 |
| Moderna | 3 | 647 | 2,256 |
| Oxford | 3 | 1,010 | 2,288 |
| Pfizer | 27 | 513 | 6,042 |
| Sputnik V | 0 | 153 | 700 |

The most common associations in COVID-19 with vaccine manufacturers indicate progress in development, rollout and are health-related (e.g. side effects). Detailed analysis of n-grams for each vaccine developer are in Supplementary Section 2. Vaccines by Moderna and Pfizer were chiefly associated with n-grams indicating progress of clinical trials and their rollouts. By contrast, top n-grams associated with AstraZeneca and Johnson & Johnson were linked to side effect reporting by these companies (unexplained illness, blood clot). Throughout the pandemic, Sputnik V, is mentioned not in medical context but rather frequently linked to Russia and Vladimir Putin, containing frequent n-grams like *“Soviet Union”, “President Vladimir Putin”, “Russia Soviet Union”*.

We investigated the extent to which the difference in context of the vaccine manufacturers influenced the sentiment of the news articles. In Figure 6 we plot the proportion of negative and positive sentiments towards the vaccine manufacturer entities in the two periods before vaccine and after vaccine. In the period before vaccine rollout, there is no entity with notably different polarization from all other entities. In the period after vaccine rollout, AstraZeneca has notably a higher ratio of negative articles and lower ratio of positive articles. Despite Johnson & Johnson also being associated with side effects (as per our n-gram analysis), AstraZeneca received notably worse press as judged by sentiment polarization. We removed AstraZeneca coverage from Figure 3 and Figure 4 to test whether its higher associated volume of negative news was influencing the slightly more negative polarization in the rollout phase. In neither case have we found that AstraZeneca was the main driver in more negatively polarized articles in that period.

**Figure 6:**
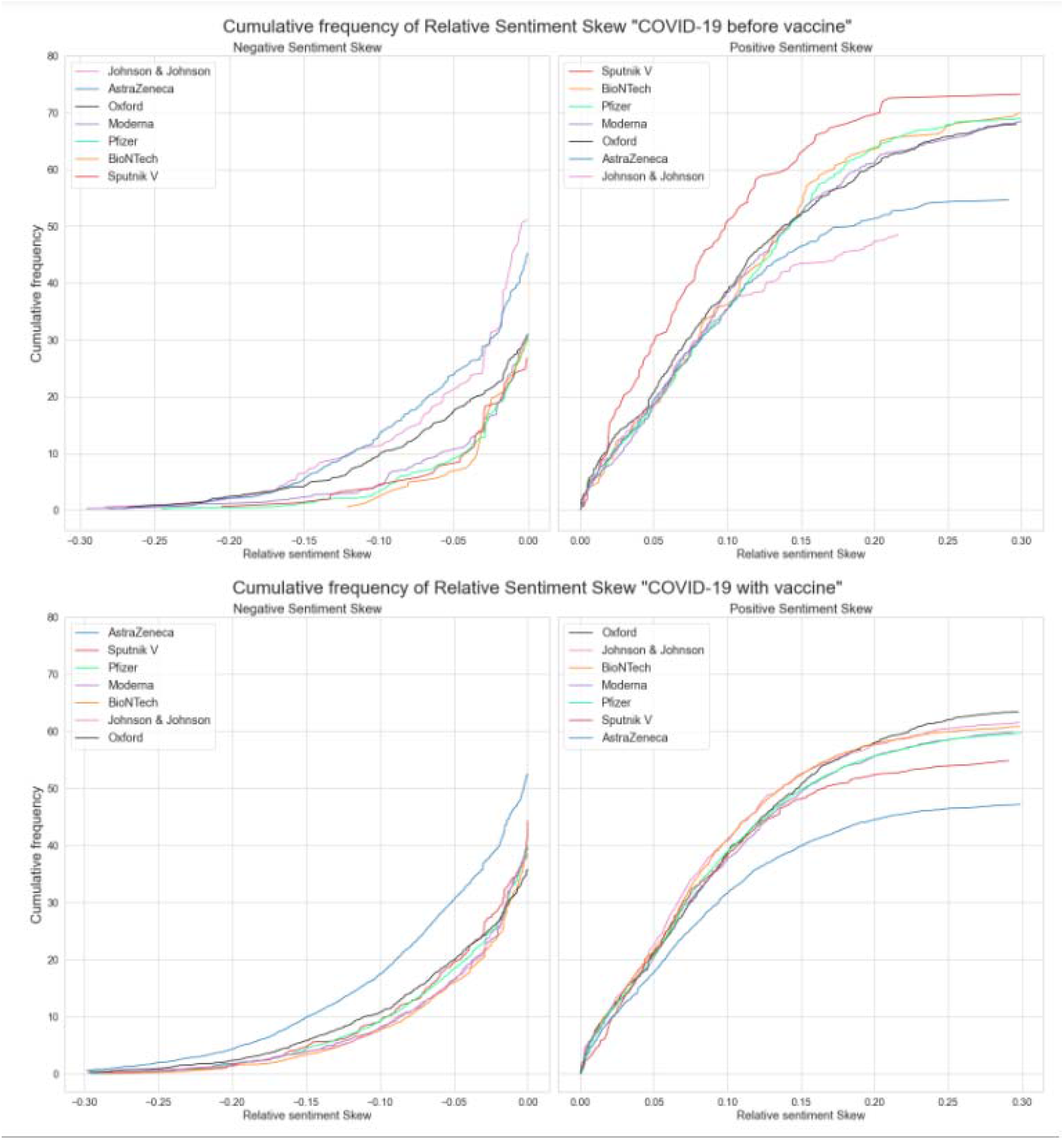
Proportion of negative and positive sentiment polarization with respect to entities associated with vaccine manufacturing in the period before vaccine rollout and after vaccine rollout.

## 4. Discussion

We used text-mining to study vaccine reporting on the front pages of top national news outlets. As willingness to receive a jab is dependent on perception of the vaccine, traditional news media is important in shaping this perception.

We analyzed the extent to which vaccine reporting increased due to the COVID-19 pandemic. For this purpose we chose a keyword-based approach that was previously used to measure the extent of COVID-19 reporting (19). The approach is designed to increase the precision of identified headlines, though at the expense of recall. For instance, the headline: *“UK measles outbreak: 500,000 British children don’t have crucial jab - Daily Star. MORE than half a million children in the UK didn’t receive a…”* was not extracted for the English vaccine dataset, as it does not contain any of the chosen key words given in in Table 1, even though it is clearly about vaccination. Developing a more complex topic model would not guarantee better performance and comparability between different languages, as one would have to develop a suitable model that captures the same linguistic nuances. Therefore, we resorted to simple mentions of basic vaccine-derived keywords.

Though our approach inherently underestimates the number of vaccine-related articles, we could confirm that as a result of COVID-19, vaccine reporting was given central prominence, and was not reported only sporadically as before the outbreak. Studying the volume of vaccine coverage motivated our choice of division of the study into the three periods, before COVID-19, during COVID-19 but before vaccine, and with COVID-19 vaccine. Specifically, the division between the second and third period could have been done in different ways, which could have influenced our results. We found it reasonable to make these divisions according to the large rise in the relative frequency in vaccine headlines, which was a result of the Pfizer and BioNTech press release on 9 November, 2020. This press release influenced all countries, while many of the other cornerstones in this period were more country specific. For instance, the UK was the first country to approve the Pfizer-BioNTech vaccine on 2 December, 2020, with USAs FDA approval of the Pfizer-BioNTech vaccine on 11 December, 2020.

Our topic modeling and sentiment analysis showed a considerable shift in both volume and sentiment of vaccine reporting. COVID-19 increased the volume of reporting on vaccines from negligible 0.1% to 3.84% during rollout across our 172 ONSs. Reporting on vaccines in the period prior to COVID-19 pandemic was majority negatively polarized. By contrast, even though in absolute terms, the number of negatively polarized vaccine articles increased several-fold, overall the vaccine-related reporting during the pandemic carries positive sentiments. This is despite well-reported side-effects associated with certain vaccines such as AstraZeneca, of which we note a reflection in the data, though not significant enough to account for the majority of negative reporting. Though Oxford co-created the vaccine, it does not experience an equally large proportion of negative headlines as AstraZeneca, which might be reflected in the media coverage frequency of the two with respect to vaccines. While AstraZeneca is mentioned 5,881 times during the pandemic, Oxford is mentioned 3,298 times, of which the majority is in COVID-19 before vaccine, while for AstraZeneca the majority is in the subsequent period. Which could indicate that AstraZeneca is more frequently connected with the vaccine in the media coverage than Oxford.

Overall our results provide a quantification of the increased vaccine reporting during the COVID-19 pandemic. How the increased coverage, though predominantly positive, will translate into public perception of vaccines is yet to be seen by traditional survey-based studies. Insofar, we hope that our study will provide a text-mining basis for possible vaccine-perception shaped by major traditional news media.

## Data Availability

Data are available via sciride.org

http://sciride.org

## Funding

This work was supported by Centre funding from the UK Medical Research Council under a concordat with the UK Department for International Development, the NIHR Health Protection Research Unit in Modelling Methodology and Community Jameel. This research was also partly funded by the Imperial College COVID-19 Research Fund. S. Bhatt acknowledges The UK Research and Innovation (MR/V038109/1), the Academy of Medical Sciences Springboard Award (SBF004/1080), The MRC (MR/R015600/1), The BMGF (OPP1197730), Imperial College Healthcare NHS TrustBRC Funding (RDA02), The Novo Nordisk Young Investigator Award (NNF20OC0059309) and The NIHR Health Protection Research Unit in Modelling Methodology.

## Notes

### Competing Interest Statement

The authors have declared no competing interest.

### Author Declarations

This study does not involve any human data.

