## Supplementary File for "Quantifying changes in vaccine coverage in mainstream media as a result of COVID-19 outbreak"

### 1. Topic Detection

#### Choosing the optimal model

The scatter plots in figures 1, 2 and 3 show how the embeddings of the headlines are scattered with respect to each other for the three different periods. The scatter in each plot are colour coded, with respect to which country they are from. Figure 1, 2 and 3 clearly show, there are topics only dealing with Australia and Canada with respect to vaccines in all three periods, while there in “COVID-19 with vaccine” also seem to be a cluster containing headlines about Ireland and vaccines. Particularly in “Before COVID-19” there does only seem to be a couple of clear clusters of headlines in the data, while there seem to be areas with more dense scatter than other areas.

Figure 2 indicates that the period “COVID-19 before vaccine” has perhaps five or seven clear clusters, depending on the minimum size of a cluster. The big multicoloured area in the middle is though characterized by being more dense in some areas, indicating that it could be a collection of several topics. This is somewhat similar with figure 3, which shows the embeddings for “COVID-19 with vaccine”, here there clearly are five clusters of headlines, which stand out from the remaining, but the big multicoloured group of headlines, is like with figures 1 and 2 characterized by having varied density within.

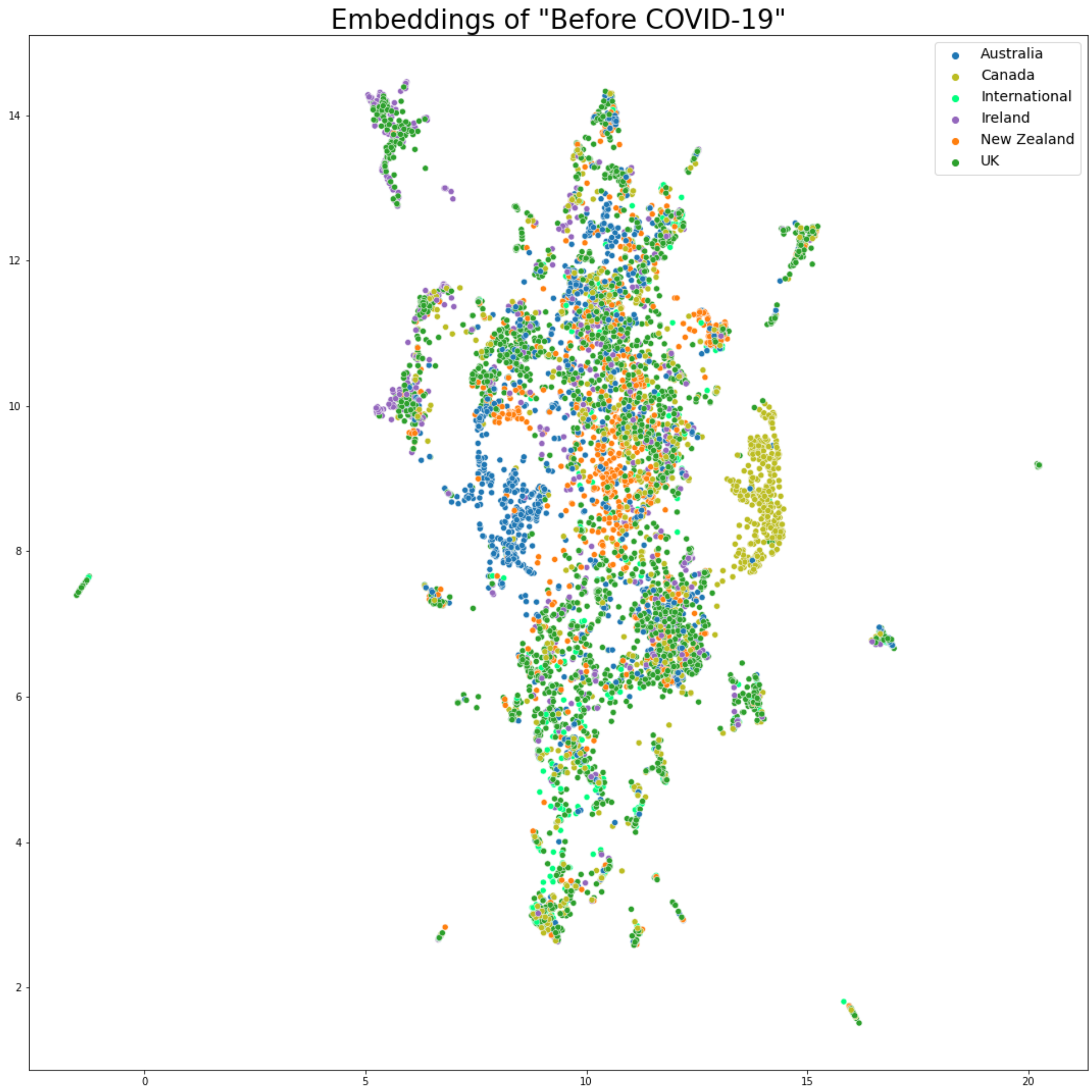


***Supplementary Figure 1**: Embedding for before COVID-19*

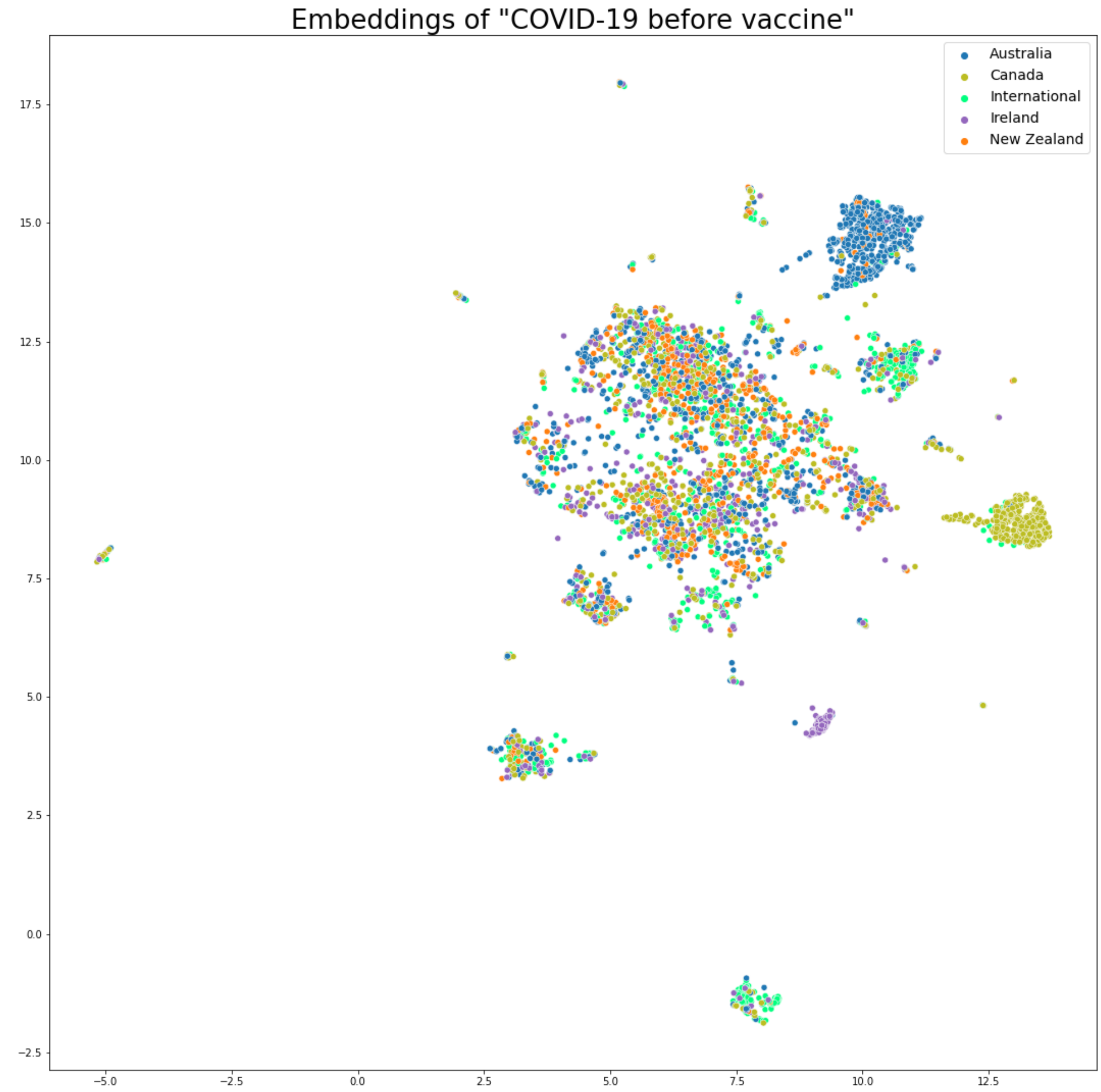


***Supplementary Figure 2**: Embedding for COVID-19 before vaccine*

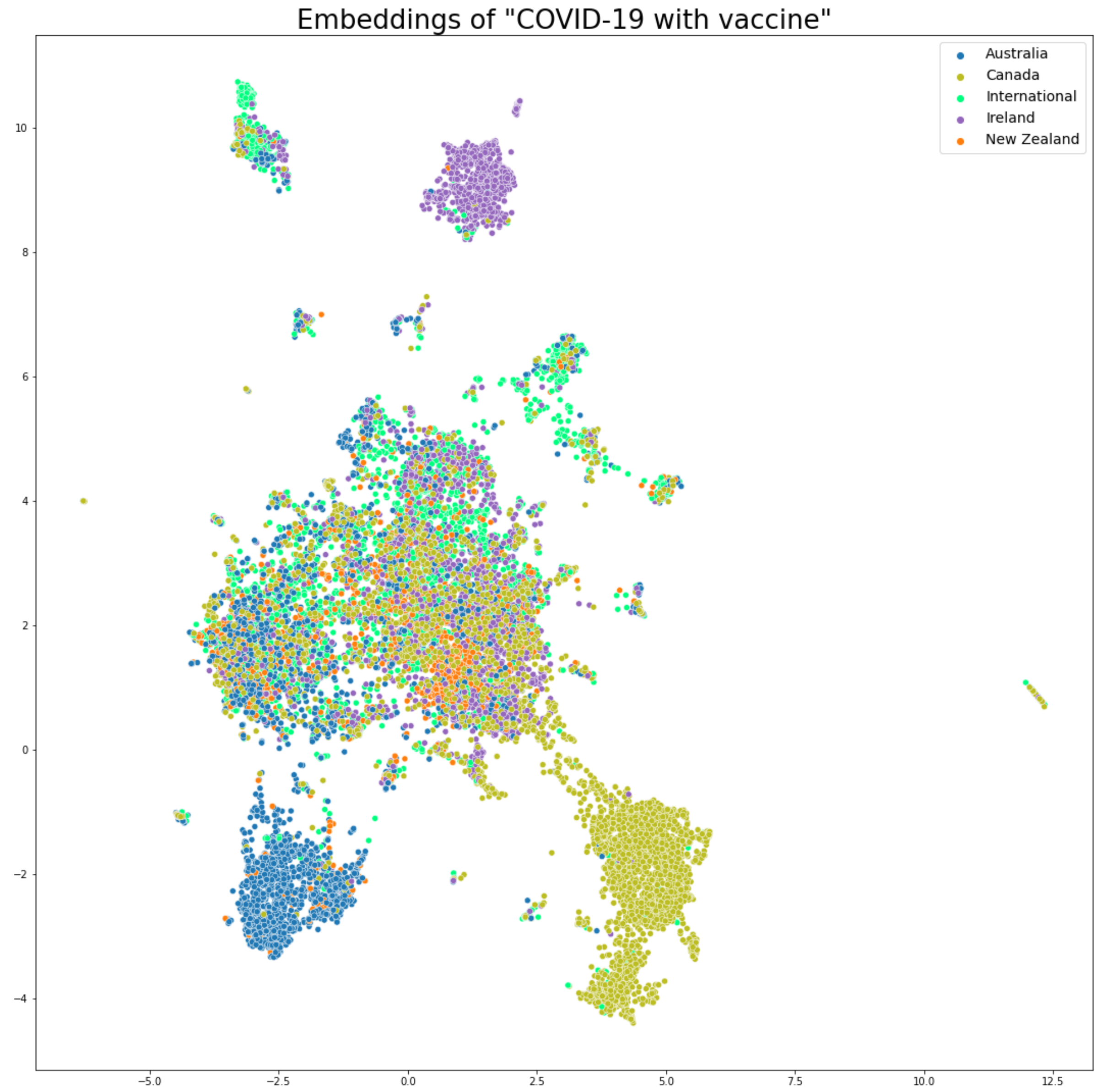


***Supplementary Figure 3****: Embedding for COVID-19 with vaccine*

### 2. Analysis of n-grams

#### Method

##### Topics for the different vaccines or vaccine manufacturers

The results from NER were used to find the most frequently occurring COVID-19 vaccine manufacturers or specific vaccines. The seven most frequently used by both English and non-English ONSs were: “AstraZeneca”, “BioNTech”, “Johnson & Johnson”, “Moderna”,

“Oxford”, “Pfizer” and “Sputnik V”. These will for simplicity be referred to as vaccines in the remainder of the report.

To learn if a possible change in sentiment from “COVID-19 before vaccine” to “COVID-19 with vaccine”, with respect to the individual vaccines, was caused by certain developments or changes over time, an assessment of the different vaccines were done for both periods. To make sure that only words used, which could help clarify, what the individual headlines were about, the lemmatized headlines, created for the topic detection, were assessed. These were additionally stemmed, to make sure that words, which only differences were their ending, would be counted as the same.

The assessment was initially done counting frequent words, but it was difficult to derive anything useful from these. Based on this frequent bigrams and trigrams were counted instead. These were derived from the headlines using “bigrams” and “trigrams” from “NLTK” [11]. These will subsequently have the common name n-grams.

For all vaccines and periods the 50 most frequent n-grams were assessed. In some cases a combination of two bigrams, with almost the same count as a trigram, would combine to that trigram. For instance the bigrams “(food, drug)” and “(drug, administr)” combined give the trigram “(food drug administr)”, where the small difference in count between bigrams and trigrams were caused, by “Food and Drug Administration” in some cases were referred to as “Food and Drug Authority” or “Food and Drug Association”. Based on this, such bigrams were removed, only keeping the trigrams. This was the case for “Food and Drug Administration”, “Centers for Disease Control” and “European Medicines Agency”. Additionally “FDA”, “CDC”, “NIH”, “WHO” and “EMA” were frequently occurring abbreviations among the frequent words with respect to some of the vaccines, whereby these were added to the number of occurrences of “Food and Drug Administration”, “Center for Disease Control”, “National Institute of Health”, “World Health Organization” and “European Medicines Authority” respectively. Other abbreviations like “NHS”, “HHS”, “PHE” etc. were assessed with respect to frequent bigrams and trigrams, to learn if they could alter the occurrence of the most frequent, which was not the case. Likewise all bigrams were assessed and if they occurred the same number of times as a trigram containing the bigram, it was removed. This was not the case if a bigram occurred more frequently than the trigram, whereby they both were kept. The bigram “(european, union)” was very frequent in most of the datasets, whereby this was removed, additionally a few ONSs were still present in the data, these were likewise removed.

As the datasets contained different amounts of headlines, it was difficult to compare the individual frequent bigrams and trigrams the vaccines and periods between. Based on this, the n-grams remaining, were used to count the number of headlines each n-grams was contained in, which was then used to calculate the given n-gram's relative frequency with respect to the number of headlines in the dataset using equation 5.1.

The 30 most frequent n-grams for each dataset were used to make a barplot, illustrating the percent of headlines each n-gram was contained in. This was done using "barplot" from "seaborn" [38].

### Results

The creation of subsets for all vaccines resulted in a total of 21 datasets, where each contained the data from "Before COVID-19", "COVID-19 before vaccine" or "COVID-19 with vaccine".

This shows a change in media coverage with respect to each period and vaccine. Sputnik V is the name of a COVID-19 vaccine, with no mention of it before the COVID-19 outbreak. The remaining vaccines are actually companies developing a COVID-19 vaccine, but there has definitely also been an increase in media attention towards these the periods between. The three periods do not have the same length, which does not matter, as the longest period is "Before COVID-19", which has the fewest observations, while the shortest is "COVID-19 with vaccine", which has the most.

The most frequent n-grams, with respect to the individual vaccines, show some similarities, while also showing some clear differences. The results for companies, which have developed their vaccine jointly, are so similar, that they will be assessed together, to not repeat the same thing several times, while the remaining will be assessed individually. Common for all vaccines is that they in "COVID-19 before vaccine" all bear a clear sign of being in a trial phase, as they all have several n-grams within the 20 most frequent, containing words like: "trial" and "test". In "COVID-19 with vaccine" all mention the "European Medicines Agency" within their 20 most frequent n-grams, while except for Sputnik V, the remaining also mention South Africa.

**AstraZeneca and Oxford** As AstraZeneca and Oxford have developed their vaccine jointly, it makes sense, if they have been mentioned in several headlines together. A count showed that they had 1901 headlines in common, whereby multiple n-grams had been counted with respect to both of them. In figures 4 and 6 it is clear, that for "COVID-19 before vaccine" AstraZeneca's and Oxford's most frequent n-grams, are heavily influenced by, the fact that they were in a trial phase, this can among other be seen in the most frequent n-gram for both of them, which occur in 8.71 percent of Oxford's headlines and 9.77 percent of AstraZeneca's. The period though also carry sign of problems with the vaccine during the trial phase, as both of them have the frequent bigrams "(trial, hold)" and "(trial, paus)", while AstraZeneca also have "(trial, resum)".

The frequent n-grams for "COVID-19 with vaccine" show that both companies experience some media attention with respect to blood clots. While blood clots are mentioned in 12.84 percent of the headlines also mentioning AstraZeneca, it is only in 5.65 percent of headlines with respect to Oxford. Which, with the difference in total headline mentioning the two, is almost 5 times as many headlines mentioning the combination AstraZeneca and blood clots than the combination Oxford and blood clots.

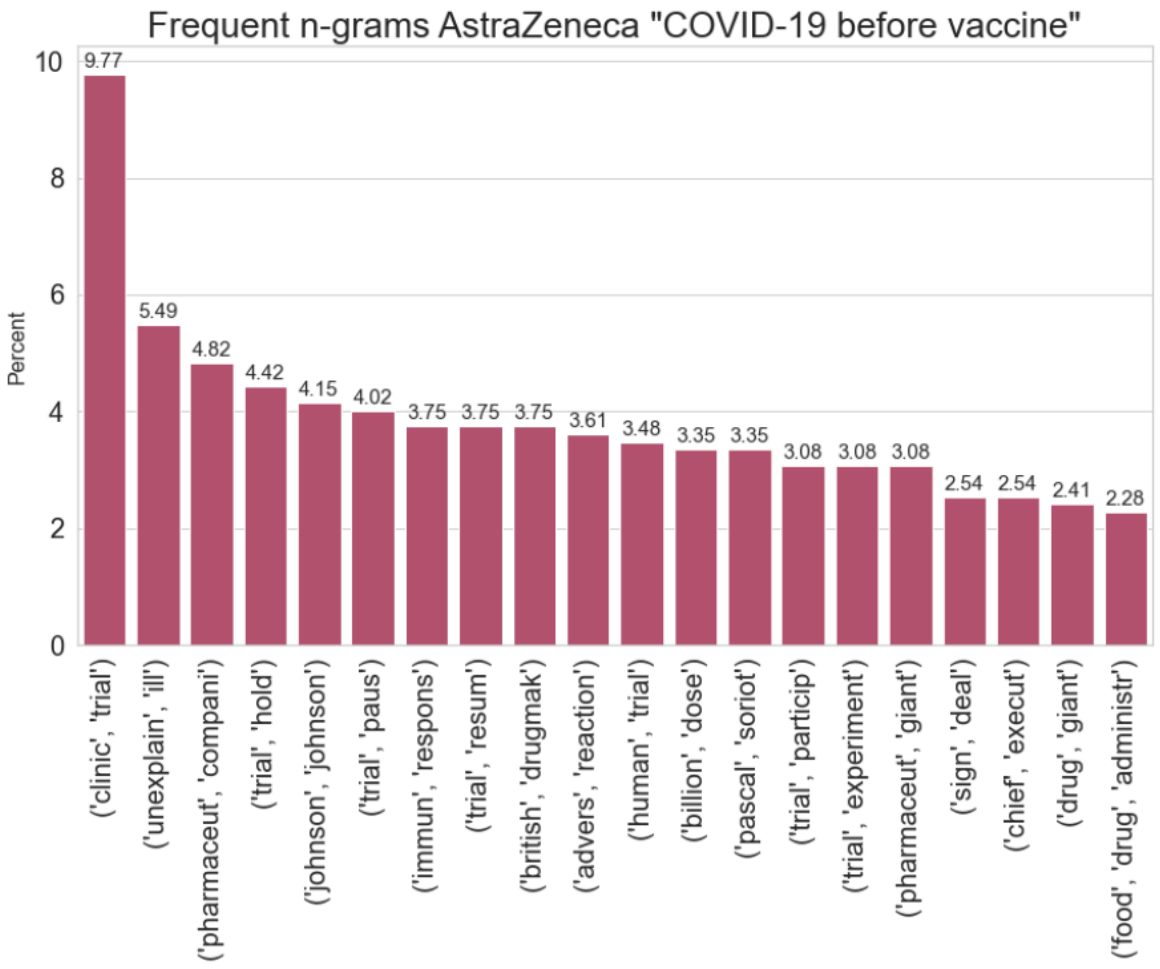


***Supplementary Figure 4:*** *The 20 most frequent bigrams and trigrams in headlines which are about AstraZeneca for the period "COVID-19 before vaccine".*

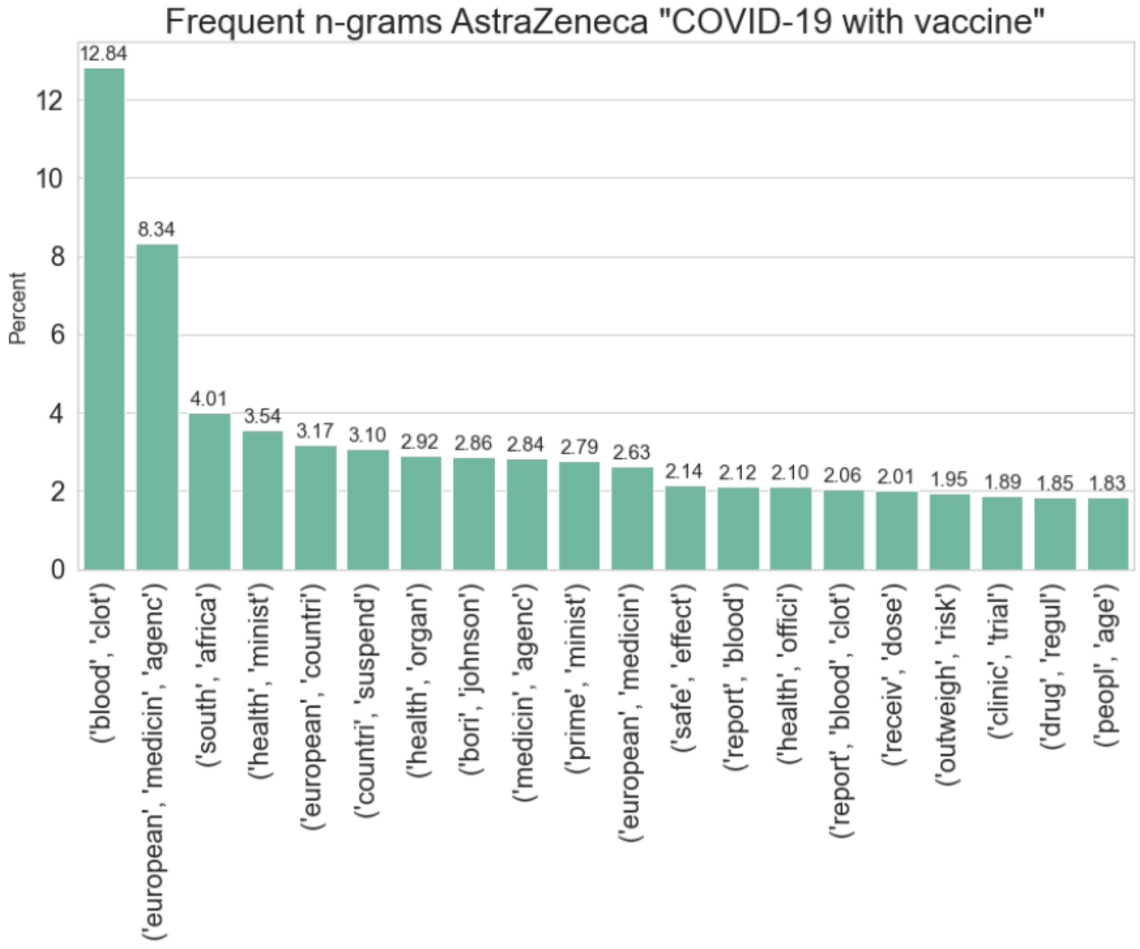


***Supplementary Figure 5:*** *The 20 most frequent bigrams and trigrams in headlines which are about AstraZeneca for the period "COVID-19 with vaccine".*

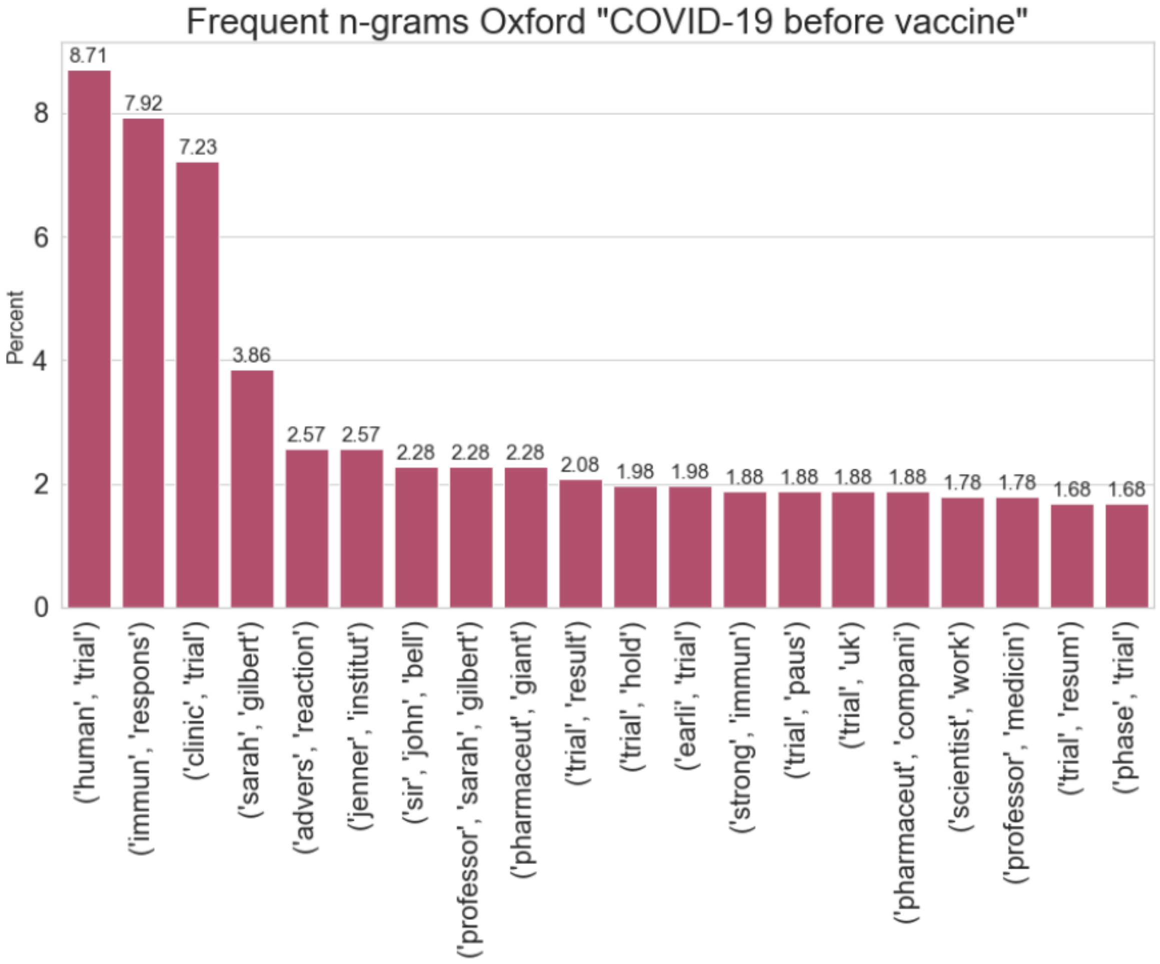


***Supplementary Figure 6:*** *The 20 most frequent bigrams and trigrams in headlines which are about Oxford for the period "COVID-19 before vaccine".*

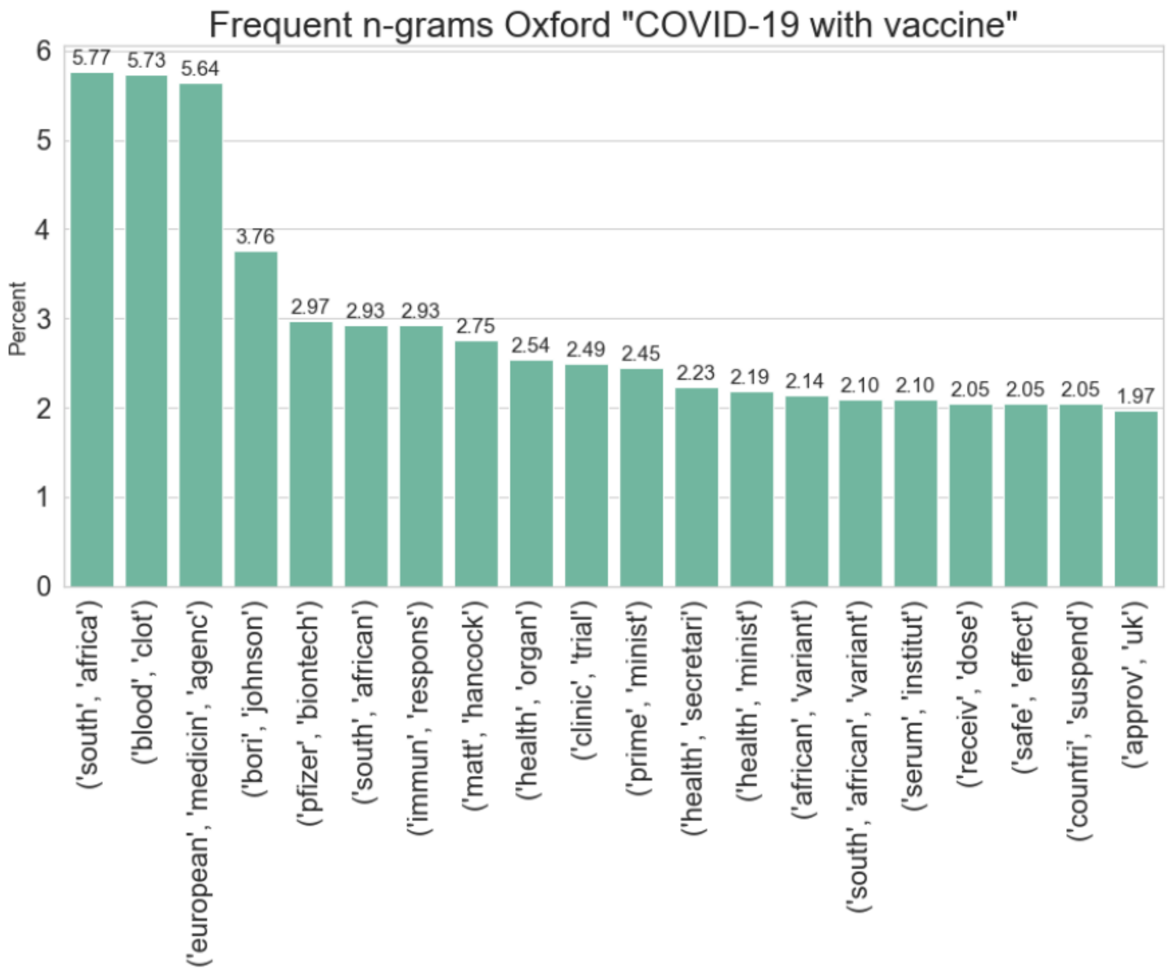


***Supplementary Figure 7:*** *The 20 most frequent bigrams and trigrams in headlines which are about Oxford for the period "COVID-19 with vaccine".*

**Johnson & Johnson** Figures 8 and 9 illustrate the frequent ngrams with respect to Johnson & Johnson. For “COVID-19 before vaccine” there seem to be two tendencies. The first is that Johnson & Johnson are in a trial phase, like all other vaccines. Additionally the period seems to carry a lot of weight with respect to unexplained illness, which caused their trial to pause. This can amongst others be seen in the bigrams: “(unexplained, ill)”, “(trial, paus)”, “(trial, halt)” etc. The unexplained illness in their study have had a lot of media attention as the bigram “(unexplain, ill)” occurs in 12.35 percent of the Johnson & Johnson headlines for this period.

In “COVID-19 with vaccine” the media have a lot of focus on the fact that Johnson & Johnson, in contrast to any of the other already approved vaccines, is a single dose vaccine, which is mentioned in 14.90 percent of the headlines.

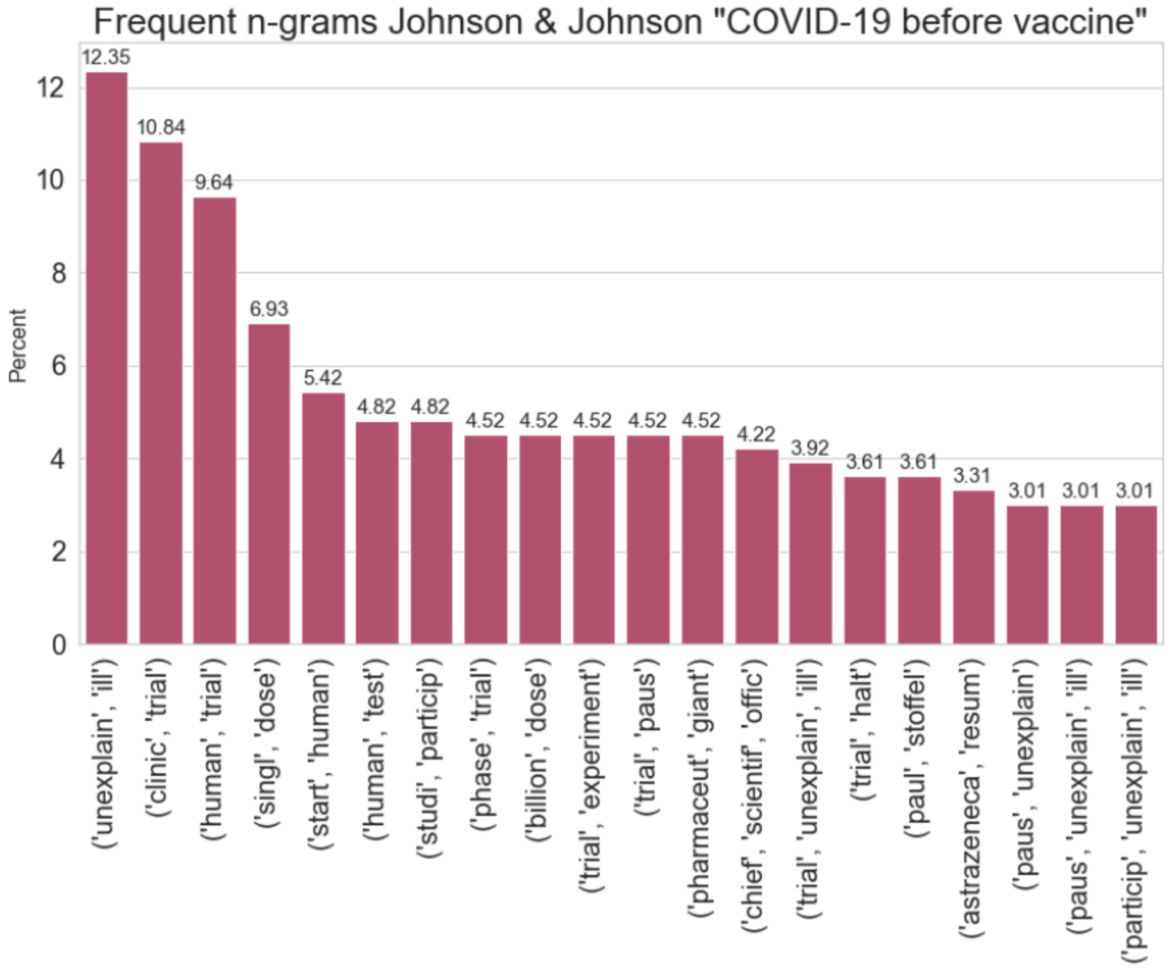


***Supplementary Figure 8:*** *The 20 most frequent bigrams and trigrams in headlines which are about Johnson & Johnson for the period "COVID-19 before vaccine".*

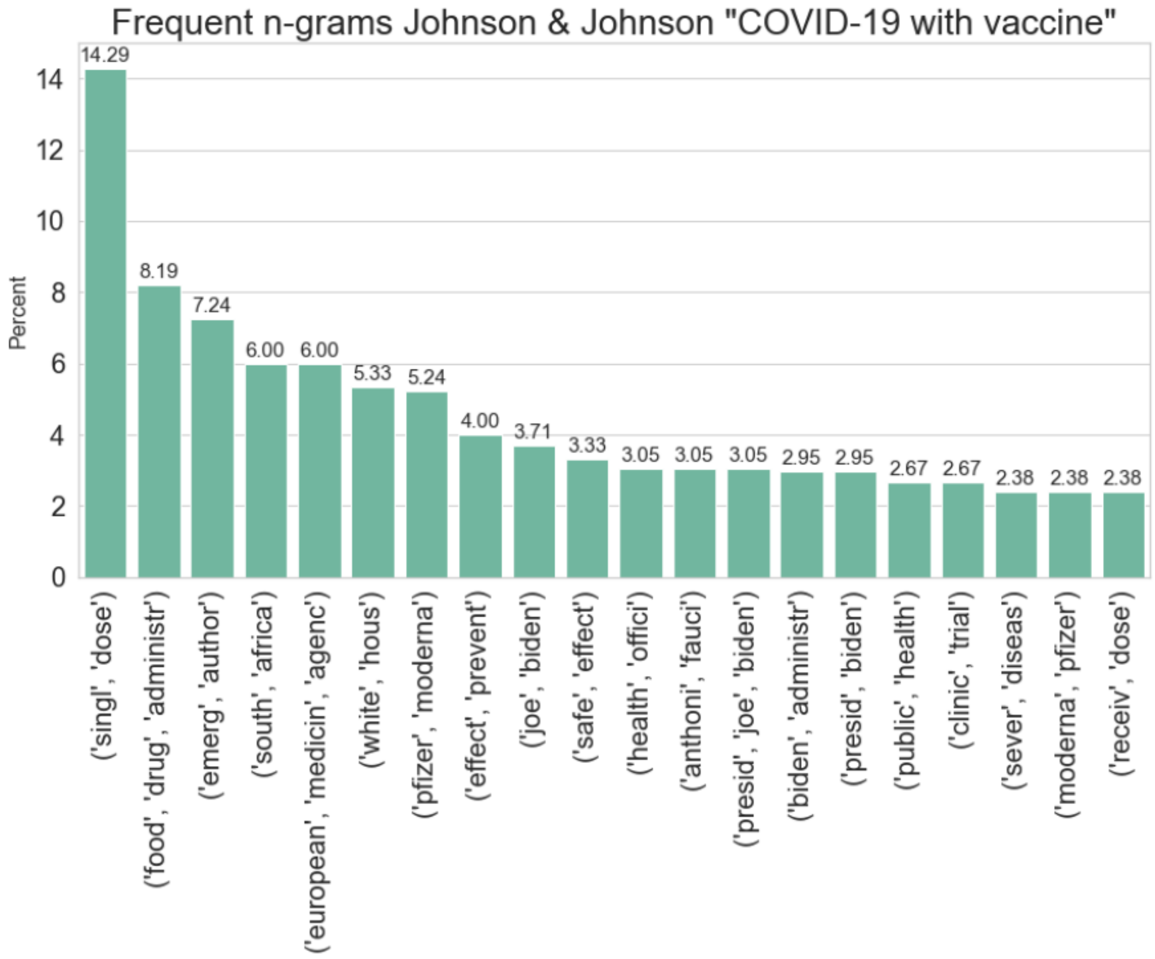


***Supplementary Figure 9:*** *The 20 most frequent bigrams and trigrams in headlines which are about Johnson & Johnson for the period "COVID-19 with vaccine".*

**Pfizer and BioNTech** Like AstraZeneca and Oxford, Pfizer and BioNTech have developed their vaccine jointly, whereby it likewise makes sense, that they have a lot of frequent n-grams, with respect to their headlines in common. A search showed that 2,125 headlines did actually mention them both.

Like all other vaccines “COVID-19 before vaccine” is characterized by being a trial period. Additionally it show signs of an upcoming approval, with n-grams like “(fast, track)”, “(fast, track, status)”, “(sign, deal)” and “(regulatory, approval)” being frequent n-grams in BioNTech, while “(emerg, author)” and “(late, novemb)” are indicators for Pfizer, this can be seen in figures 10 and 12.

In “COVID-19 with vaccine” Pfizer and BioNTech both have relative low frequency with respect to their most frequent n-grams, where the two most frequent for both of them are “(european, medicin, agenc)” and “(receiv, dose)”, which for Pfizer occur in 4.94 percent and 4.08 percent of the headlines respectively, while in BioNTech’s headlines they occur in 7.11

percent and 4.82 percent respectively. In this period there does not seem to be any particular subject, which is in focus, with respect to the media for any of the two vaccines.

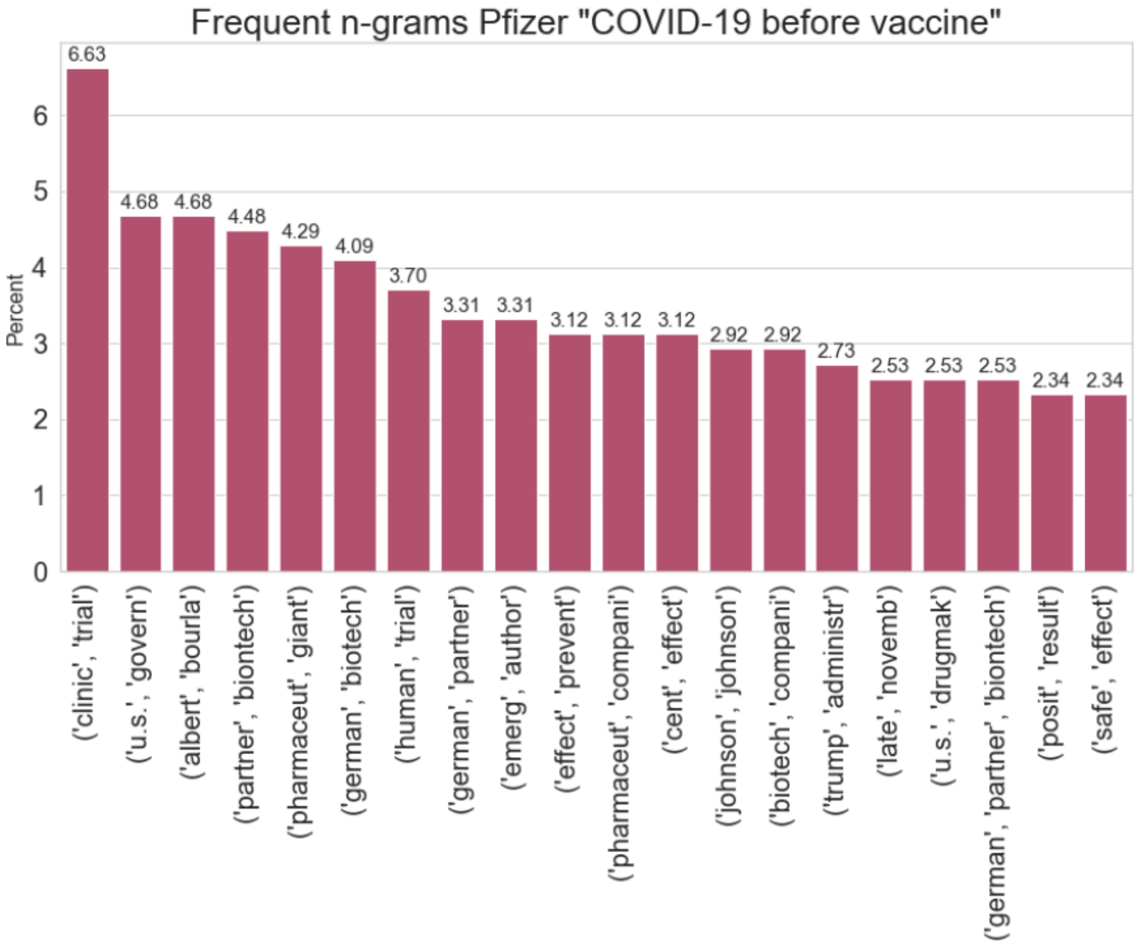


***Supplementary Figure 10:*** *The 20 most frequent bigrams and trigrams in headlines which are about Pfizer for the period "COVID-19 before vaccine".*

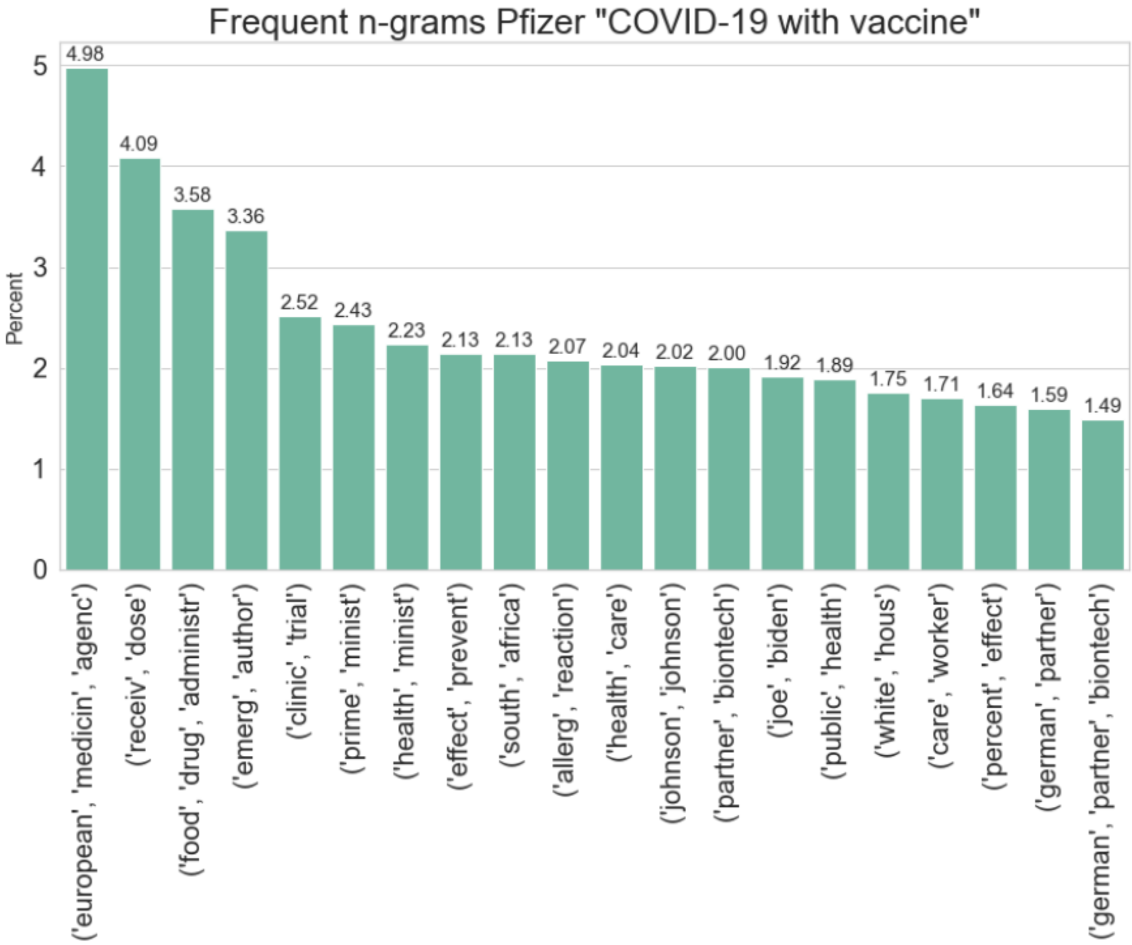


***Supplementary Figure 11:*** *The 20 most frequent bigrams and trigrams in headlines which are about Pfizer for the period "COVID-19 with vaccine".*

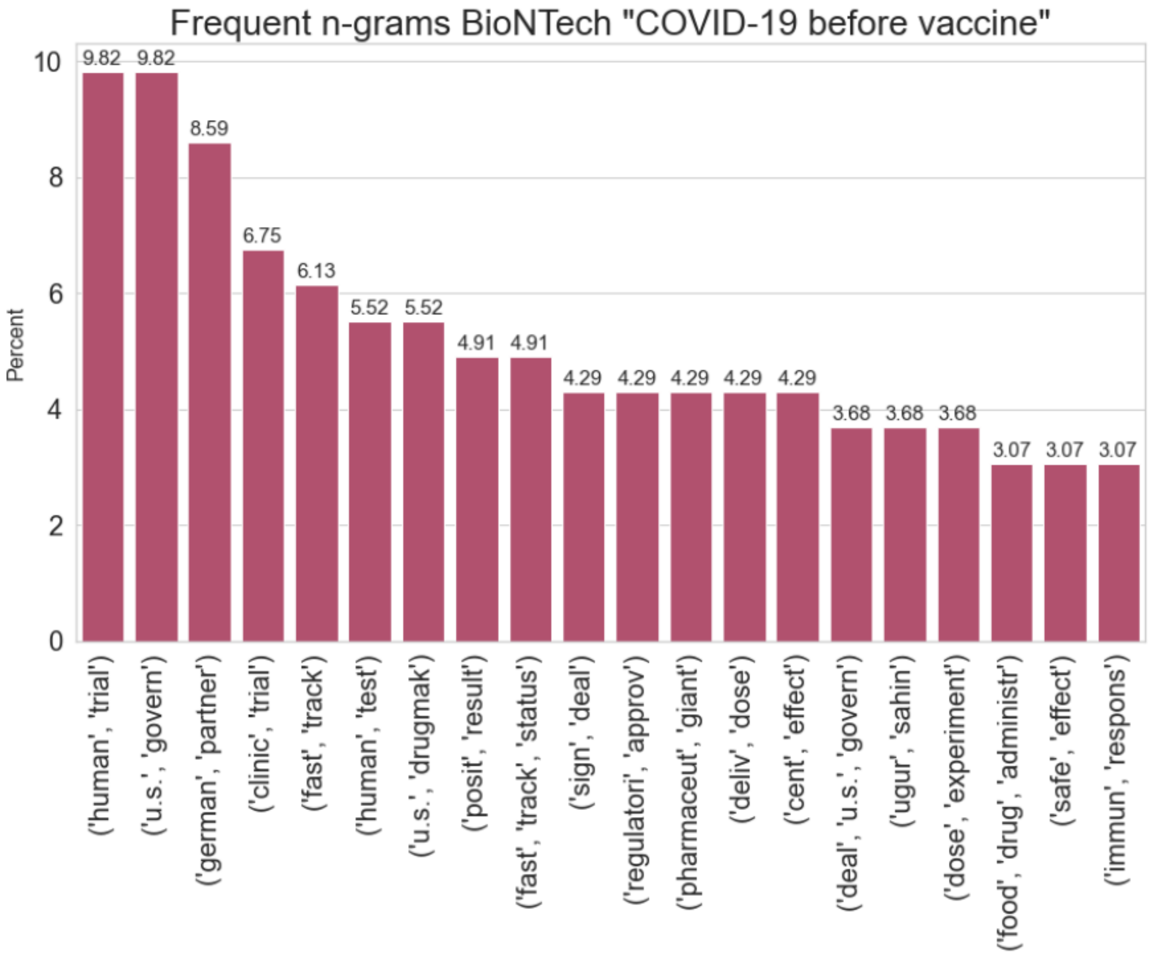


***Supplementary Figure 12:*** *The 20 most frequent bigrams and trigrams in headlines which are about BioNTech for the period "COVID-19 before vaccine".*

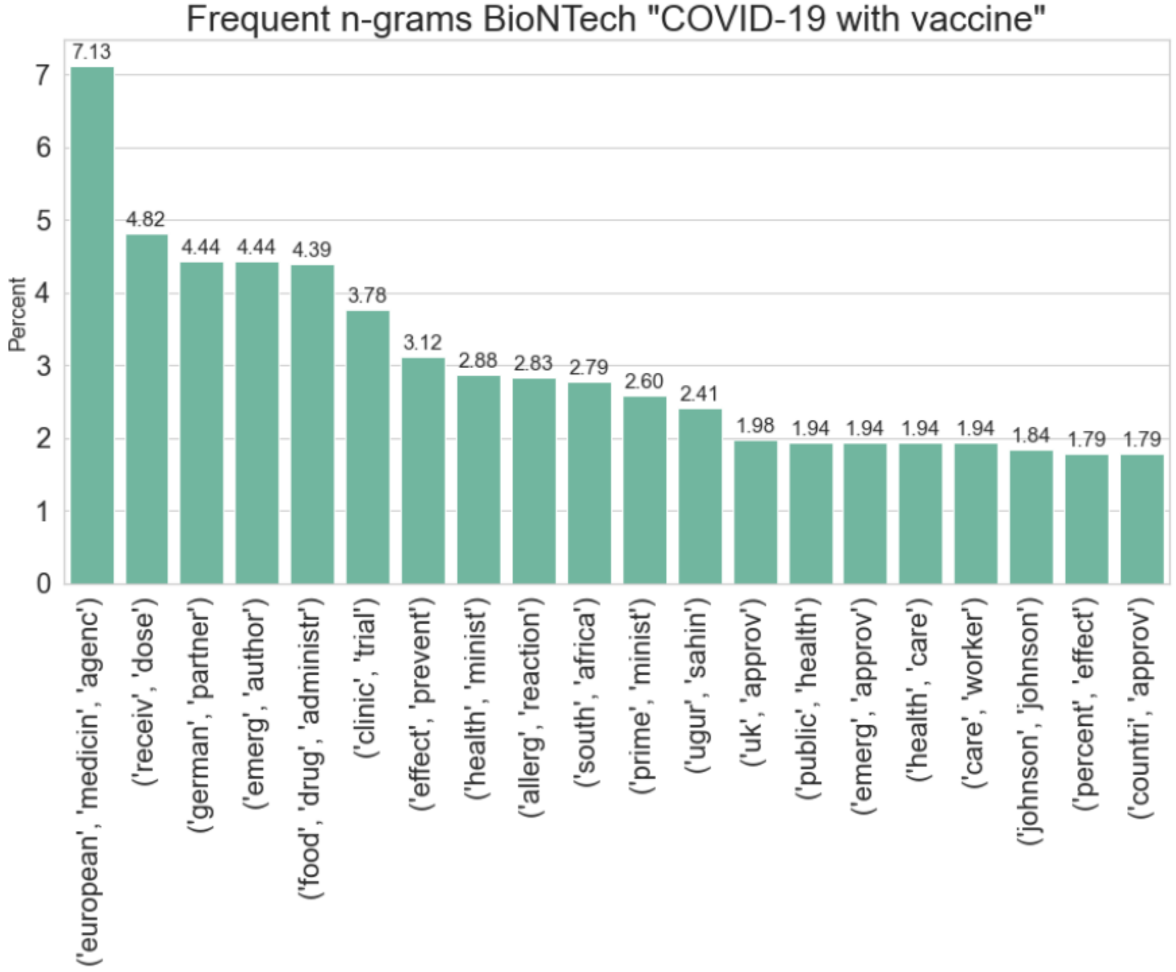


***Supplementary Figure 13:*** *The 20 most frequent bigrams and trigrams in headlines which are about BioNTech for the period "COVID-19 with vaccine".*

**Moderna** In “COVID-19 before vaccine” Moderna does not show any sign of other subjects getting some high media attention with respect to the frequent n-grams, other than being in a trial phase, this can be seen in the 20 most frequent n-grams, illustrated in figure 14. This also shows that in 3.25 percent of the headlines, Moderna is mentioned in combination with “(nation, institut, health)”, whereby it is the only vaccine, having NIH among its most frequent ngrams.

For “COVID-19 with vaccine” the bigram “(pficer, biontech)” is the most frequent, occurring in 9.29 percent of the headlines. Additionally it is the only vaccine having “(center, diseas, control)” among its 20 most frequent n-grams. 2.27 percent of the headlines in this period also mention Dolly Parton, which probably is caused by her being one of their major funders.

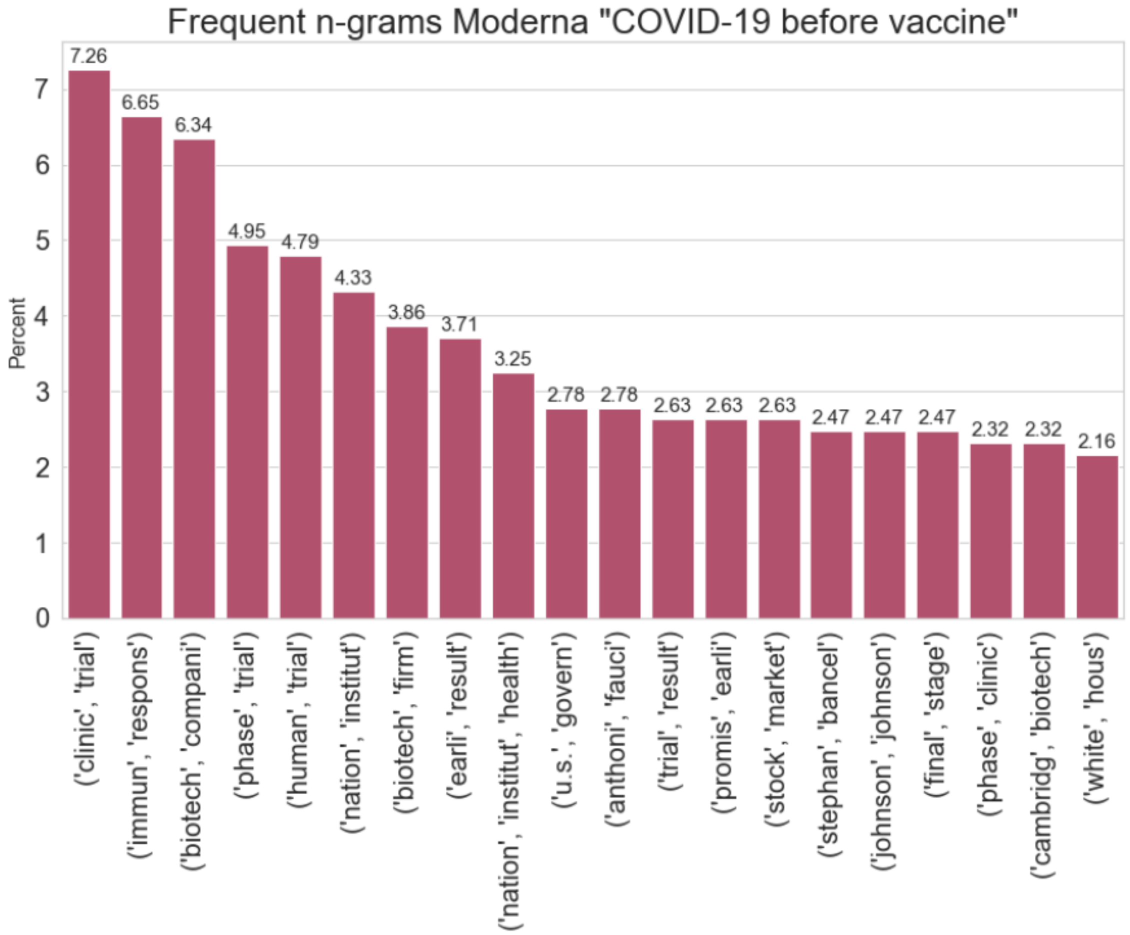


***Supplementary Figure 14:*** *The 20 most frequent bigrams and trigrams in headlines which are about Moderna for the period "COVID-19 before vaccine".*

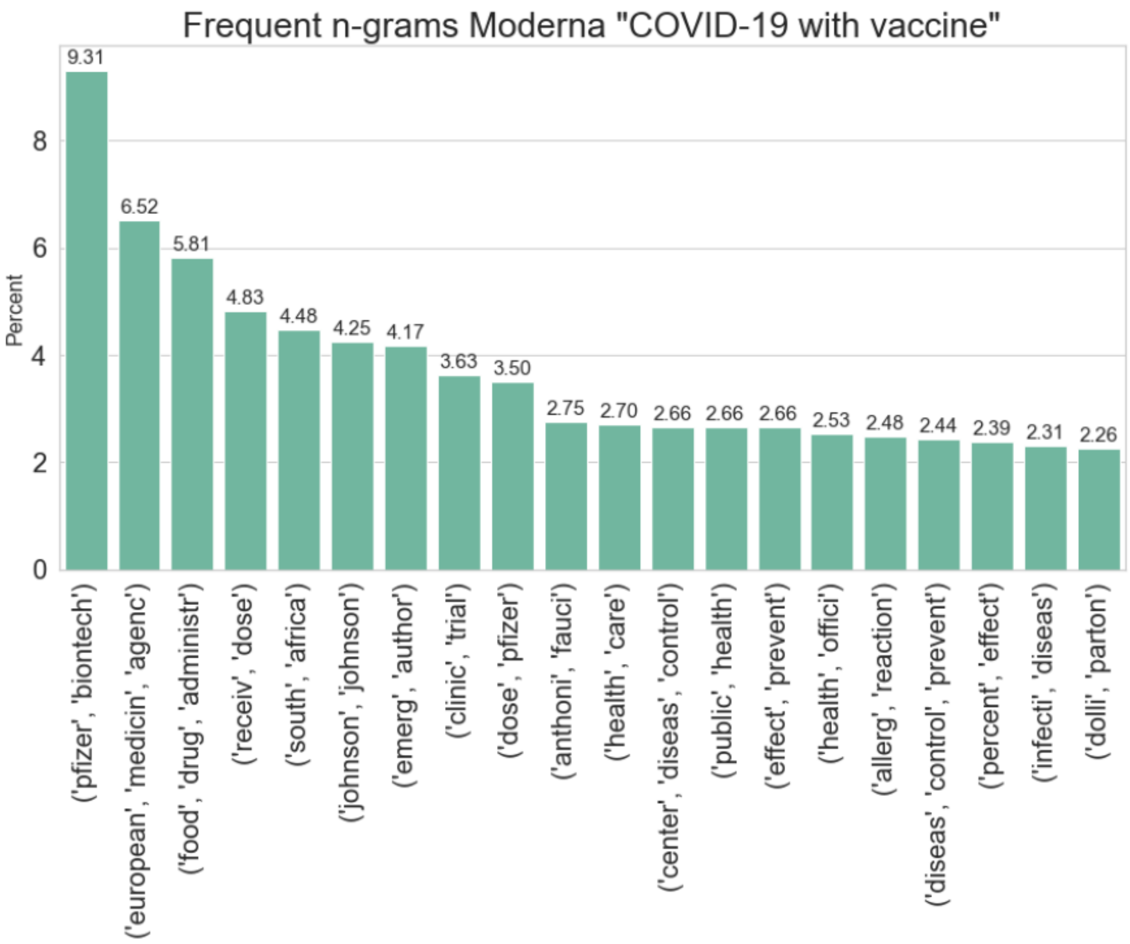


***Supplementary Figure 15:*** *The 20 most frequent bigrams and trigrams in headlines which are about Moderna for the period "COVID-19 with vaccine".*

**Sputnik V** In figures 16 and 17 the 20 most frequent n-grams with respect to Sputnik V for the two periods are illustrated. In contrast to any of the other vaccines, the headlines in both periods have clear indications of the vaccine's origin, in this case Russia. For instance the most frequent n-gram in “COVID-19 before vaccine”, with a relative frequency of 31.37 percent, is “(soviet,union)”, while the second highest, with almost as many headlines, is a trigram containing the most frequent bigram, namely “(russia, soviet, union)”. Apart from the clear sign of country of origin, Sputnik V in “COVID-19 before vaccine”, like the other vaccines, have signs of being in a trial period, with the bigrams “(trial, russia)” and “(clinic, trial)” occurring in 11.11 percent and 9.15 percent of the headlines respectively.

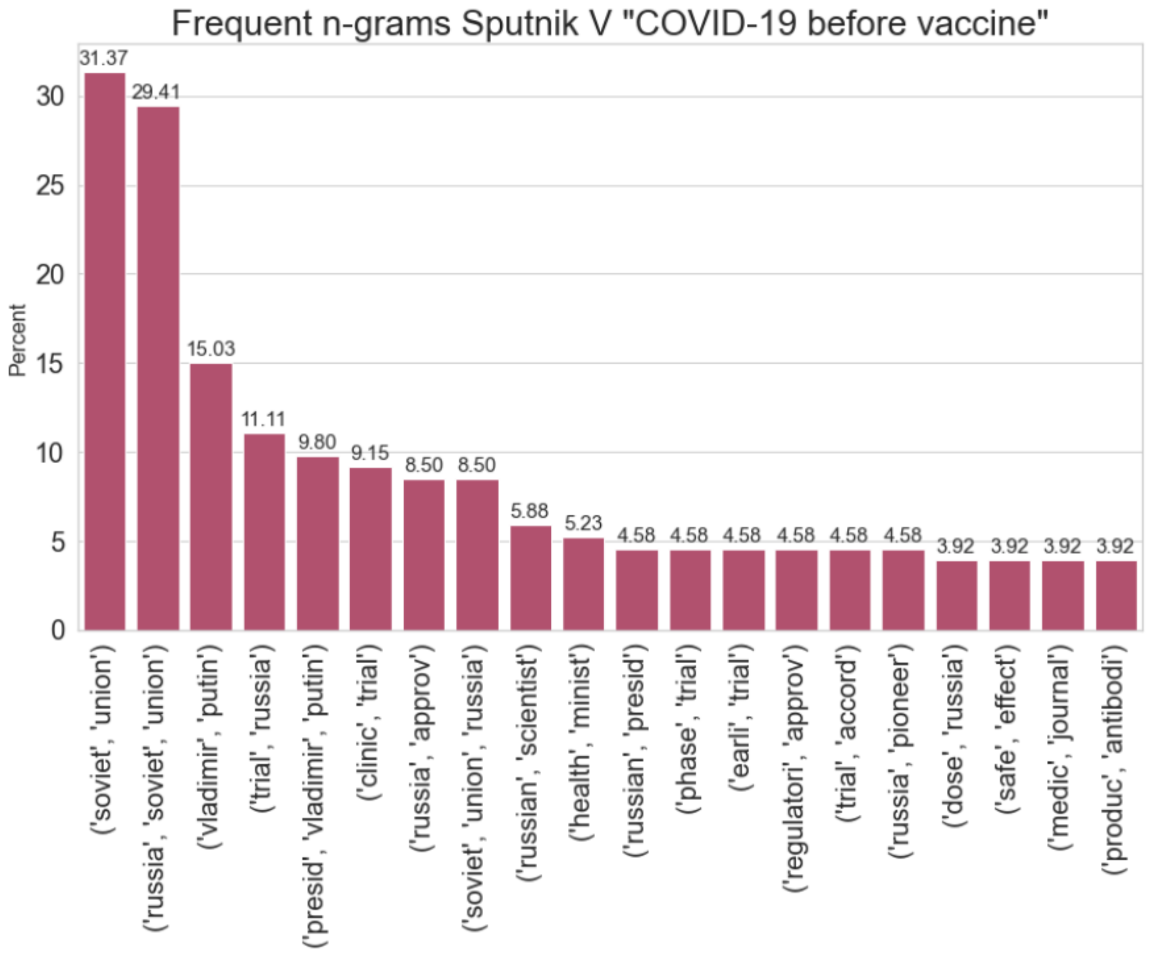


***Supplementary Figure 16:*** *The 20 most frequent bigrams and trigrams in headlines which are about Sputnik for the period "COVID-19 before vaccine".*

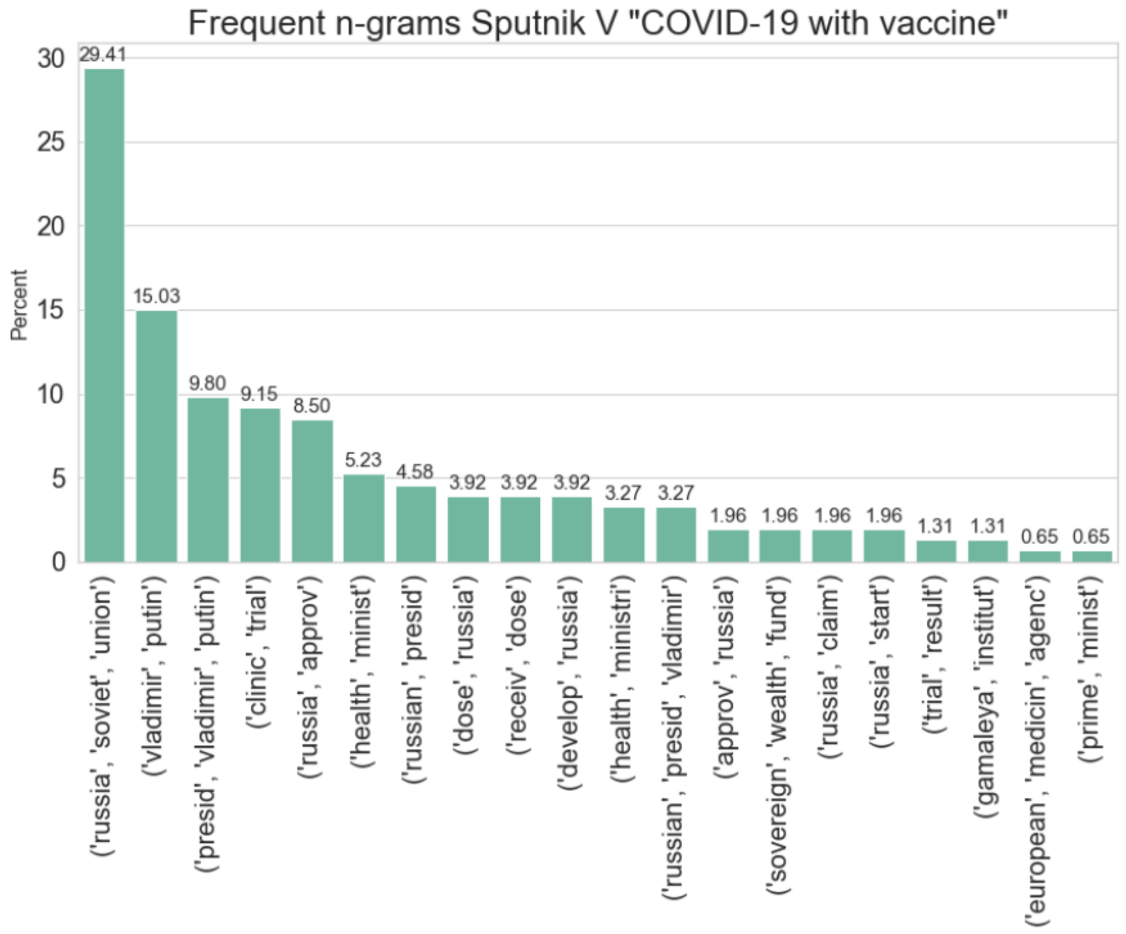


***Supplementary Figure 17:*** *The 20 most frequent bigrams and trigrams in headlines which are about Sputnik for the period "COVID-19 with vaccine".*

The frequent n-grams for “COVID-19 with vaccine” illustrate, that removing some of the Russian n-grams, to achieve a more nuanced collection of n-grams in both periods, would not have achieved much, as 9 of the 20 most frequent n-grams for “COVID-19 with vaccine”, occur in less than 2 percent of the headlines, with 3 of these being in less than 1 percent. The removal could have been defended with the fact that many of the n-grams for Sputnik V in both periods are somehow repetitions of the same phrases, like with the two most frequent n-grams in “COVID-19 before vaccine”. A look at the 50 most frequent n-grams with respect to “COVID-19 before vaccine”, actually showed that the pattern with “Russia” and “trial” related n-grams continue being frequently mentioned after the first 20 illustrated in figure 16.
